# Genome-wide association analysis of composite sleep health scores in 413,904 individuals

**DOI:** 10.1101/2024.02.02.24302211

**Authors:** Matthew O Goodman, Tariq Faquih, Valentina Paz, Pavithra Nagarajan, Jacqueline M Lane, Brian Spitzer, Matthew Maher, Joon Chung, Brian E Cade, Shaun M Purcell, Xiaofeng Zhu, Raymond Noordam, Andrew J. K. Phillips, Simon D. Kyle, Kai Spiegelhalder, Michael N Weedon, Deborah A. Lawlor, Jerome I Rotter, Kent D Taylor, Carmen R Isasi, Tamar Sofer, Hassan S Dashti, Martin K Rutter, Susan Redline, Richa Saxena, Heming Wang

**Author notes:** Corresponding author: Heming Wang, PhD, Division of Sleep and Circadian Disorders, Brigham and Women’s Hospital, Harvard Medical School, 221 Longwood Ave BLI 252, Boston, MA 02115.

## Abstract

Recent genome-wide association studies (GWASs) of several individual sleep traits have identified hundreds of genetic loci, suggesting diverse mechanisms. Moreover, sleep traits are moderately correlated, and together may provide a more complete picture of sleep health, while also illuminating distinct domains. Here we construct novel sleep health scores (SHSs) incorporating five core self-report measures: sleep duration, insomnia symptoms, chronotype, snoring, and daytime sleepiness, using additive (SHS-ADD) and five principal components-based (SHS-PCs) approaches. GWASs of these six SHSs identify 28 significant novel loci adjusting for multiple testing on six traits (p<8.3e-9), along with 341 previously reported loci (p<5e-08). The heritability of the first three SHS-PCs equals or exceeds that of SHS-ADD (SNP-h^2^=0.094), while revealing sleep-domain-specific genetic discoveries. Significant loci enrich in multiple brain tissues and in metabolic and neuronal pathways. Post GWAS analyses uncover novel genetic mechanisms underlying sleep health and reveal connections to behavioral, psychological, and cardiometabolic traits.

## INTRODUCTION

Sleep is an essential biological process, orchestrated by interrelated neurologic and physiologic regulatory processes, responding to individual, social, and environmental influences^1–3^. Positive sleep traits have been associated with lower rates of cardiometabolic and neuropsychiatric diseases, as well as higher productivity and well-being^4^. Moreover, general sleep health has come into recent focus as a consequential and modifiable health factor, with the combined presence of multiple healthy sleep factors frequently being a stronger predictor of positive health outcomes^5,6^. As a composite, sleep health is recognized to involve multiple domains, including regularity, satisfaction, alertness, timing, efficiency, and duration^1^.

Several recent studies leveraging biobank-scale data have resulted in well-powered genome-wide association studies (GWASs) of sleep phenotypes, capturing several aspects of sleep health, including self-reported sleep duration^7^, insomnia^8,9^, sleepiness^10^, snoring^11^, and chronotype^12^. These GWASs have begun to elucidate the genetic architecture of sleep, while revealing the presence of widespread genetic correlations across both sleep and related neuropsychiatric and cardiometabolic traits^2,3^. Associated genomic loci and pathways are often shared across multiple sleep traits, suggesting a shared genetic basis and co-regulated processes. Therefore, a more complete and robust understanding of sleep may be achieved by describing patterns across multiple traits, pointing toward underlying domains, highlighting the potential utility in analyzing composite sleep health scores (SHSs).

Recently, an additive sleep health score (SHS-ADD) consisting of five self-reported sleep behaviors was studied in unrelated individuals in the UK Biobank (UKB), yielding new genetic findings^13^. However, additive scores compress data across multiple domains to a single metric, resulting in potential information loss, increased genetic heterogeneity, and weaker signal for genetic analyses. In this study, we expanded the prior UKB SHS-ADD GWAS to a larger UKB sample and constructed five novel principal-component (PC)-derived SHSs as linear combinations of the same five underlying traits. We conjectured that, when compared with SHS-ADD, SHS-PCs would create more precisely targeted phenotypes, resulting in potentially greater heritability, as well as distinct and interpretable domain-specific associations in secondary analyses.

## RESULTS

### Sleep health score construction in UKB

The study population consisted of 413,904 UKB participants of European ancestry with complete sleep and genomics data (Methods). Sample characteristics are provided in Supplementary Table 1. Self-reported sleep traits (sleep duration, insomnia, chronotype, snoring, and daytime sleepiness) derived from the baseline questionnaire were used to construct SHS traits (Methods). Briefly, SHS-ADD was operationalized as in previous UKB studies^13,14^, defined as the sum of five dichotomized positive sleep health characteristics: sleep duration of 7 to 8 hours, morning chronotype preference, no snoring, infrequent insomnia symptoms and infrequent daytime sleepiness (Methods). SHS-PCs (mean-centered and variance-standardized) were extracted from the same underlying sleep traits, treated as linear continuous measures, and then oriented to positively correlate with self-assessed overall health.

Several findings emerged from constructing the Sleep Health Score Principal Components (SHS-PCs) in the UK Biobank, providing context and guiding interpretation. SHS-PCs 1-5 individually explained from 25.2% to 14.9% of the phenotypic variation (Fig. 1 and Supplementary Table 2). Based on their PC loadings (Fig. 1 and Supplementary Table 2), higher scores on SHS-PCs are interpreted as follows – SHS-PC1: longer sleep with less-frequent insomnia symptoms and sleepiness; SHS-PC2: healthier sleep with less-frequent sleepiness and without snoring (i.e., without symptoms of sleep apnea syndrome); SHS-PC3: morningness chronotype; SHS-PC4: snoring with less frequent sleepiness; SHS-PC5: shorter sleep duration with less-frequent insomnia symptoms. Substantial non-normality was observed for SHS-PC2, SHS-PC3, and SHS-PC4 (Supplementary Fig. 1), resulting from the underlying trait distributions (Methods). The direction and loadings of the PCs follow from underlying correlations among the self-report sleep traits (Supplementary Table 3): SHS-PC1 was driven primarily by correlations between sleep duration and insomnia (-0.24) and between insomnia and sleepiness (0.09); SHS-PC2 was driven by the correlation between sleepiness and snoring (0.08); SHS-PC3 loaded on chronotype, which was independent of the other underlying traits; whereas SHS-PC4 and SHS-PC5 appear to be driven largely by the PC independence constraint, such that they loaded on both positive and negative sleep attributes, and does not imply these combinations constitute clusters in the data.

**Figure 1.**
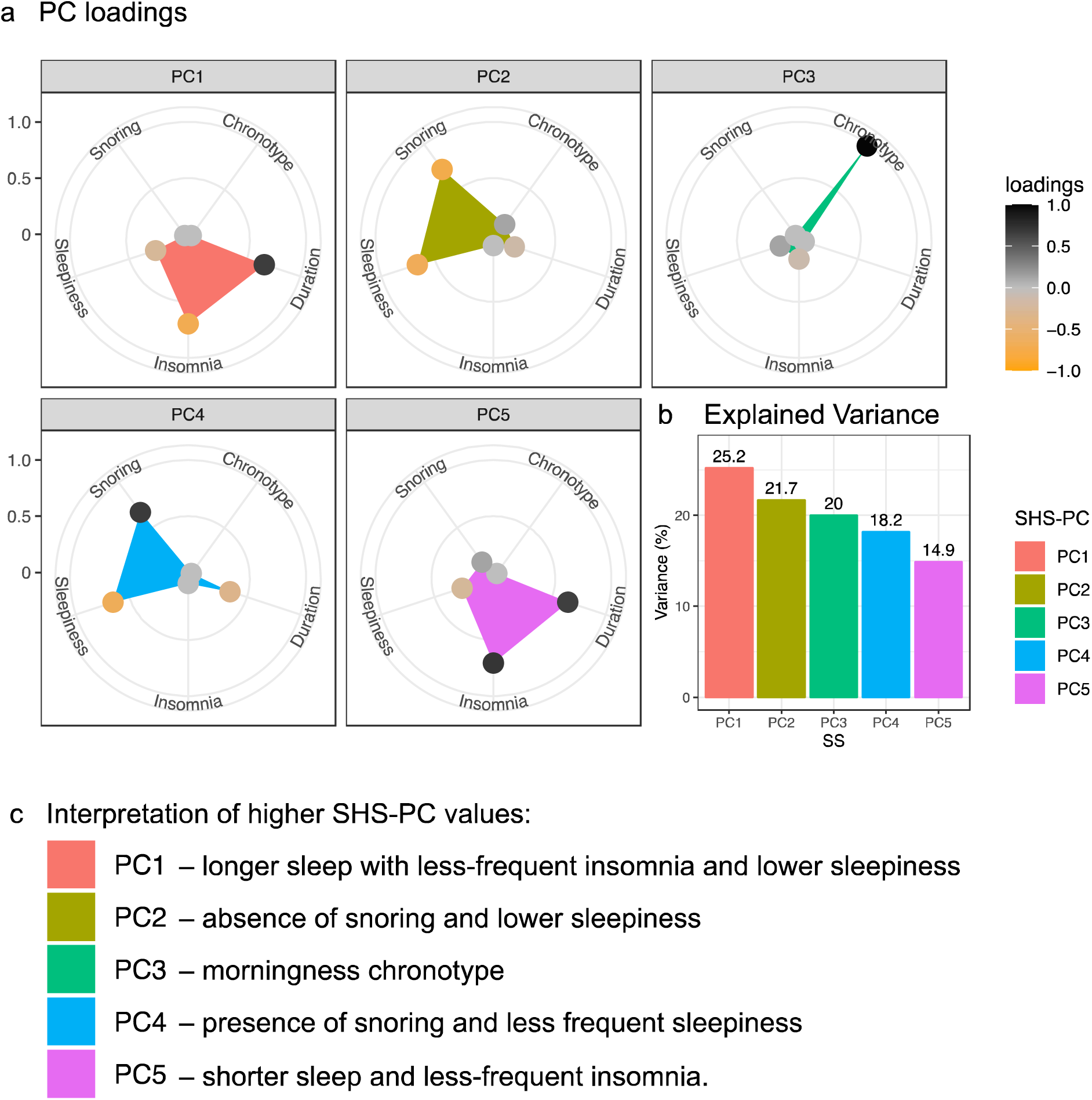
PC loadings and variance explained for the five principal component-based sleep health scores. **a.** Radar plots of loading magnitudes (radial distance). Black dots: positive loadings; Orange dots: negative loadings. **b.** Percent of the phenotypic variance explained by each sleep health score. c. Interpretations of SHS-PCs based on their loadings.

The interpretation of the SHSs was further clarified by their correlations with objective traits not used in their construction (Supplementary Figs. 2a and 3a). For example, SHS-PC1 was positively correlated with accelerometry-based sleep duration and sleep efficiency metrics. SHS-PC2 was positively correlated with higher sleep efficiency, lower daytime inactivity, lower BMI, and being female, features which suggest the absence of sleep apnea. SHS-PC3 correlated with earlier accelerometry-based measures of activity and sleep midpoint (measurements of timing).

Notably, SHS-PC4 and SHS-PC5 showed weaker correlations with self-assessed overall health (p>0.05 for PC4) but were nonetheless correlated with other measures of overall health, including lower levels of self-reported disability and lower numbers of treatments/medications taken. Notably, SHS-ADD had the strongest association with self-reported overall health, as well as with the accelerometry-derived sleep regularity index.

### Genome-wide association analysis

We performed GWAS for the six SHS traits using linear mixed regression models, adjusting for age, sex, genotyping array, ten genetic PCs and genetic relatedness matrix (Methods). We identified 31,188 genome-wide significant (GWS) SNPs (p<5e-8), resulting in 45 loci for SHS-ADD (SNP-h^2^=0.094), and 91, 48, 166, 26, and 25 loci for SHS-PC1-5 (SNP-h^2^=0.117, 0.093, 0.153, 0.070, and 0.068), respectively (Fig. 2a, Table 1, Supplementary Figs 4-5, and Supplementary Tables 4-5; Methods). Function annotation of all SNPs in linkage disequilibrium (LD; *r*^2^ ≥ 0.6) with the lead SNPs in the risk loci were performed using FUMA^15^ (Methods; Fig. 2c-2f).

**Figure 2:**
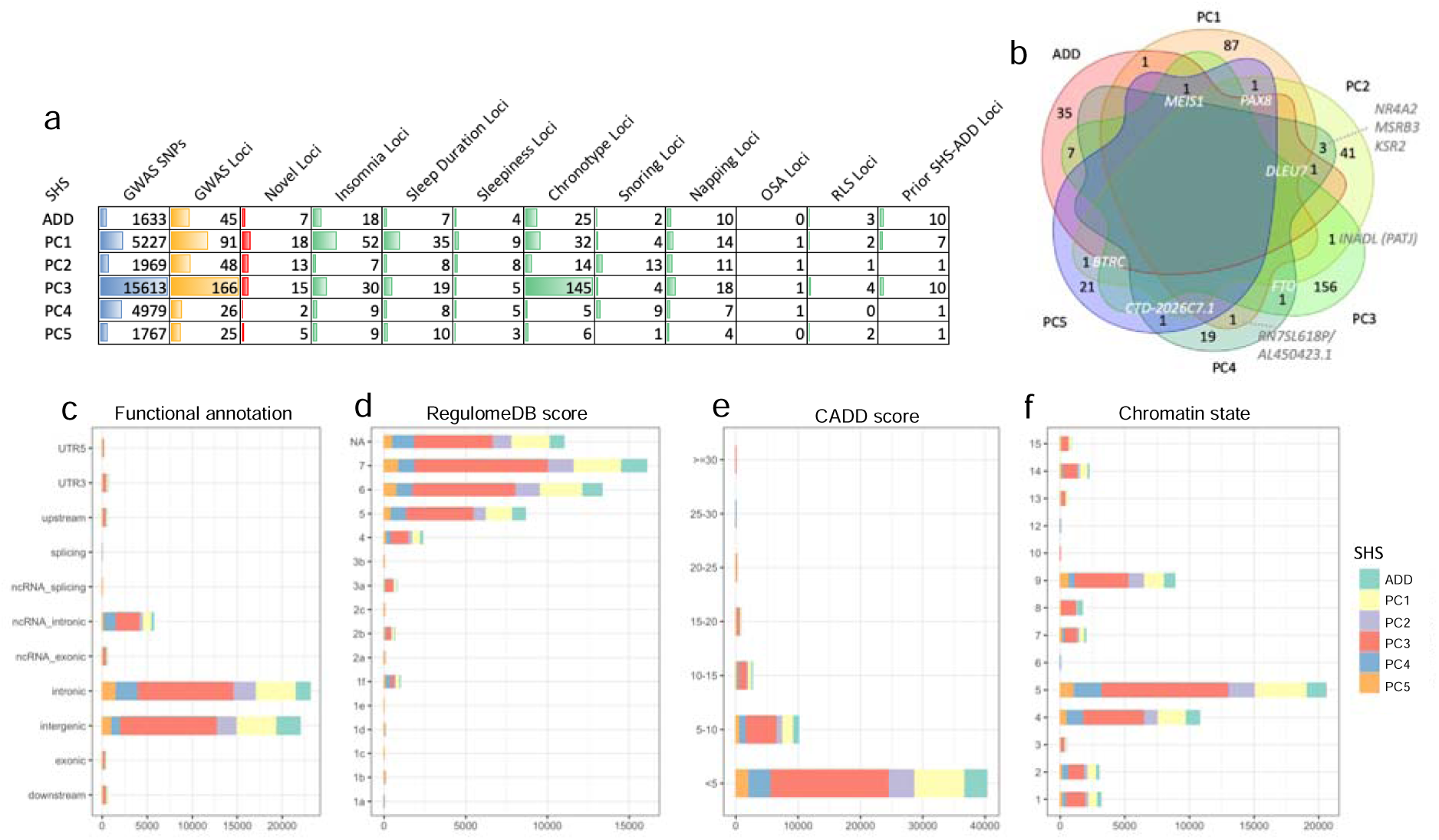
Genome-wide significant SNPs associated with SHS. **a,** Genome-wide significant SNPs and loci. Blue: Number of GWS SNPs (p<5e-08); Yellow: Number of GWS loci; Red: Number of loci not reported (at least 500kb away) by previous sleep GWASs in biobanks; Green: Number of loci reported (within 500kb) by previous sleep GWASs in biobanks. OSA: obstructive sleep apnea; RLS: restless leg syndrome. **b,** Venn diagram of GWS loci shared across SHS traits based on colocalization analysis (Supplementary Table 8). Rs113851554 at *MEIS1* (colocalizing SHS-ADD, SHS-PC1, SHS-PC3, and SHS-PC5) was reported in GWASs of insomnia, chronotype, and restless leg syndrome. Rs2863957 at *PAX8* (colocalizing SHS-PC1, SHS-PC2, and SHS-PC5) was reported for sleep duration and insomnia. Rs1421085 at *FTO* (colocalizing SHS-PC3 and SHS-PC4), a widely recognized obesity gene, was reported for sleep duration, chronotype, snoring, and OSA. **c,** Distribution of Functional consequences of all annotated SNPs in LD with independent GWS SNPs by SHS. 3.2% of the annotated SNPs were in functional regions (exon, UTR, and splice site). **d,** Regulome DB score distribution of all annotated SNPs in LD with independent GWS SNPs by SHS. 3.4% of the annotated SNPs were in regulatory regions with Regulome DB score<2. **e,** CADD score distribution of all annotated SNPs in LD with independent GWS SNPs by SHS. 6.8% of the annotated SNPs likely deleterious effect with CADD score>10. **f,** Chromatin state distribution of all annotated SNPs in in LD with independent GWS SNPs by SHS.74% of the annotated SNPs were in open chromatin regions with a minimum chromatin state between 1 and 7.

**Table 1:**
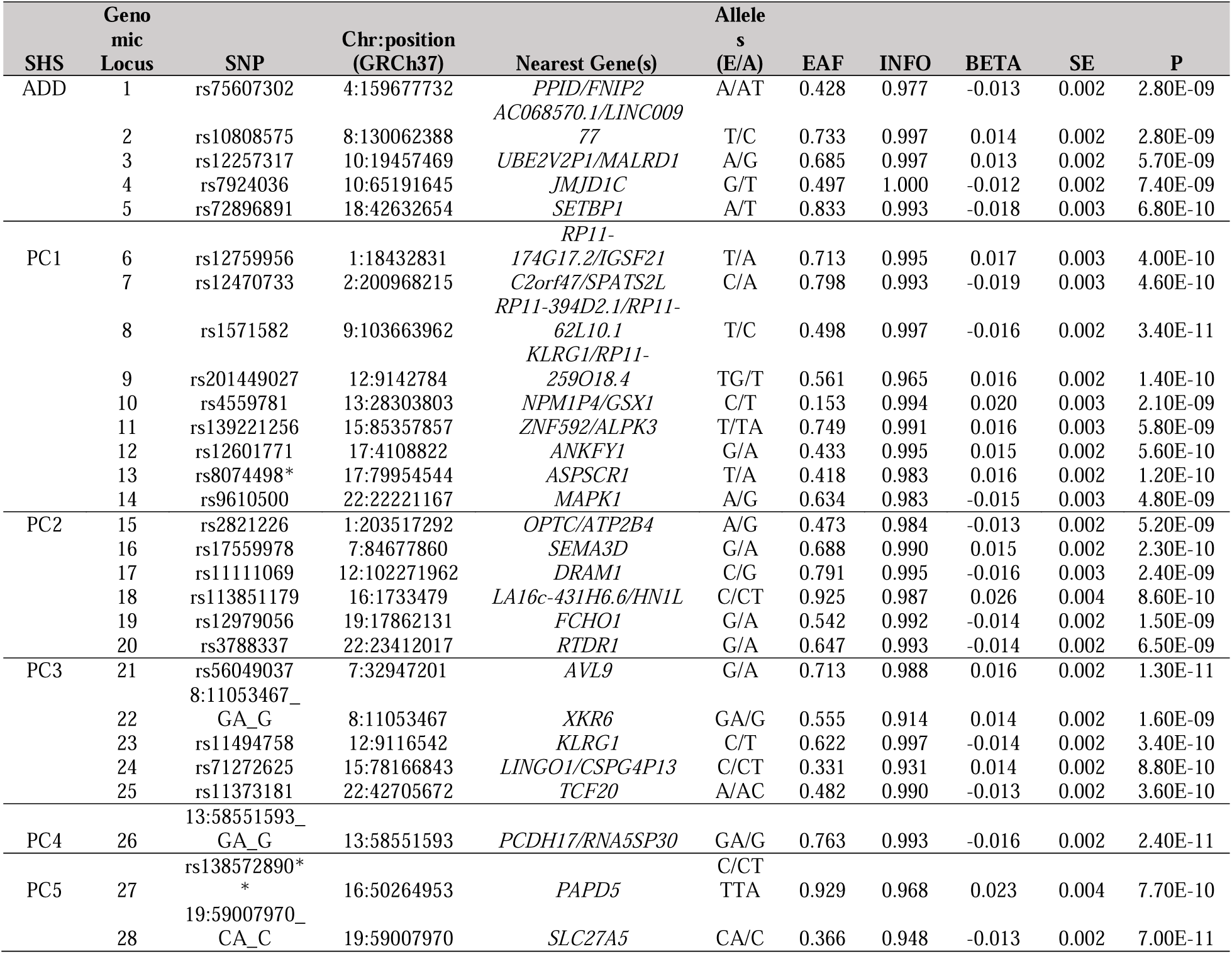
Novel loci associated with SHS (p<8.3e-9).

To determine novelty, we compared the 400 unique GWS loci against those found to be GWS in prior biobank GWASs of individual sleep traits and the previously developed SHS-ADD in the UKB (Methods; Supplementary Table 5). This identified 59 unreported SHS-associated GWS loci with a lead variant at least 500kb away from previously reported sleep variants (Table 1 and Supplementary Table 4). Of these unreported loci, 28 passed a stricter significance threshold (p<8.3e-9) accounting for multiple testing on six traits, which are defined as novel loci (Table 1). Among the 28 novel loci, two were independent (*r*^2^<0.1) but within 50kb of one another: rs201449027 (associated with SHS-PC1) and rs11494758 (with SHS-PC3), both in the *KLRG1* locus (a gene with reported immune system function^16^). The other 26 loci were distinctly associated with one SHS trait. In addition, there were two functional variants among the lead SNPs: First, SHS-PC1 associated with rs8074498, a missense variant in *ASPSCR1,* a gene regulating *GLUT4* in glucose sequestration and transportation in response to insulin. Second, SHS-PC5 associated with rs138572890 in the 3’ UTR of *PAPD5,* a non-canonical poly(A) polymerase involved in the surveillance and degradation of aberrant RNAs, including the glucose transporter *GLUT1*. Functional annotation for the lead SNPs at the 28 novel and 31 additional unreported loci is provided in Supplementary Tables 6 and 7.

Among all 400 GWS loci, there were 62 loci colocalizing SHS traits, including 18 loci colocalizing multiple SHS-PCs, suggesting the presence of instances of pleiotropy across SHS-PC traits (Methods; Fig. 2b and Supplementary Table 8). Examples include the established *MEIS1*, *PAX8*, and *FTO* loci, as well as a novel locus containing *TNFRSF14*, a gene involved in T-cell activation and signaling. The infrequent colocalization of loci associated with SHS-PC traits is consistent with their being largely genetic and phenotypically statistically independent (Supplementary Figs. 2b and 3b).

### Genetic overlap with individual sleep traits

Genetic correlations, between SHS traits with individual sleep and accelerometry traits, were qualitatively similar to the analogous phenotypic correlations described above, while often stronger (Supplementary Fig. 2). GWS SHS loci and their corresponding genetic risk scores (GRS; Methods) were associated with underlying self-reported and accelerometry derived sleep traits, largely in keeping with expectation, based on the construction of the SHS traits (Supplementary Tables 9 and 10). Conversely, approximately 50% of 1,039 GWS loci previously reported for SHS-ADD or individual sleep traits were associated with one or more SHS traits (p<5e-8; Supplementary Tables 11 and 12).

### Sensitivity analysis

We performed 22 distinct sensitivity analyses (Methods), for each of the 400 GWS loci across the six SHS traits in unrelated individuals (n=308,902), adjusting for various factors or restricting to one of three subsets (males-only, n=145,186; females-only, n=163,716; healthy-only, n=115,297). Specific covariate adjustments led to modest average attenuation (<15%) in SHS genetic effects across the GWS loci (Supplementary Table 13 and Supplementary Fig. 8). For example, adjustment of adiposity measures in SHS-PC2 (9.3%) and SHS-ADD (7.9%), mood variables in SHS-ADD (14.1%) and SHS-PC1 (9.6%), and in a healthy subset without chronic diseases in SHS-SHS-PC1 (13.5%). Nearly all individual loci remained GWS with similar effect size (standard error [SE] change<2) after additional adjustment.

### Replication and validation analyses

GWS SHS loci were not replicated in the HCHS/SOL study (n=11,144), with large ascertainment, age, population, and sample size differences, after accounting for multiple comparisons (Methods; Supplementary Tables 14 and 15). To further validate our findings, for each SHS we constructed polygenic risk scores (PRS) using genome-wide summary statistics and examined their associations with sleep phecodes in the MGB biobank (Methods; Supplementary Table 16). The PRSs of both SHS-ADD and SHS-PC1 were associated with lower odds of 6 of the 13 sleep phecodes: insomnia, obstructive sleep apnea, restless legs syndrome, sleep disorders (unspecified), organic or persistent insomnia, and sleep apnea (unspecified). SHS-PC2, SHS-PC4, and SHS-PC5 were associated with 2 sleep phecodes: obstructive sleep apnea and sleep apnea. Whereas the SHS-PC4 PRS was associated with higher odds for sleep apnea disorders (likely reflecting its positive association with snoring), increases in SHS-PC2 and SHS-PC5 were associated with lower odds for sleep apnea.

### Implicated genes

We prioritized the genes at GWS loci using three mapping methods (position, eQTL, and Chromatin Interaction [CI]) as well as MAGMA^17^ positional gene-based analysis in FUMA^15^ (Methods; Supplementary Tables 17-20; for gene-based GWAS Manhattan and QQ plots see Supplementary Figs 6-7). Hundreds of genes were implicated, of which we report only those with consistent evidence across all mapping methods, due to space considerations. The five novel SHS-ADD loci were mapped to 39 genes, with five genes in two loci supported by all four mapping methods. The latter included *FNIP2*, which binds to AMP-activated protein kinase (AMPK) and plays a crucial role in mTORC1 signaling and the regulation of heat shock protein-90 (Hsp90)^18^, which has been previously linked to sleep homeostasis and behavioral rhythms^19,20^; *NRBF2,* which is involved in circadian rhythm via control of autophagy, and nutrient and cellular homeostasis^21^; and *JMJD1C,* which plays a role in DNA repair^22^ and has been associated with Rett syndrome (OMIM 312750), which co-occurs with epilepsy and sleep disturbance^23^. Nine novel SHS-PC1 loci were mapped to 140 genes, with five genes in three loci mapped by all four methods. Of these, *NMB* encodes the neuromedin B neuropeptide linked to the endocrine and exocrine systems, body temperature, and blood pressure^24^, while at the same locus, *WDR73* is highly expressed in cerebellar Purkinje neurons, and *ZNF592* has been implicated in cerebellar atrophy; *ANKFY1* is also involved in the maintenance of cerebellar Purkinje cells that play a role in sleep-wake regulation^25,26^; *MAPK1* is part of the mitogen-activated protein kinase (MAPK)/extracellular signal-regulated kinase (ERK) signaling pathway that is linked to mental health and the circadian system^27^. Five novel SHS-PC2 loci were mapped to 97 genes (nine genes in four loci mapped by all methods). Of these, *SEMA3D* encodes a member of the class III semaphorins that are involved in axon guidance during neuronal development^28^ (two more class III semaphorins at this locus, *SEMA3A* and *SEMA3E*, also mapped by CI); *DRAM1* is a regulator of autophagy in the context of mitochondrial dysfunction, implicated in neurodegeneration^29^; *MAPK8IP3* is another MAPK/ERK gene essential for the function and maintenance of neurons, with links to neurodevelopmental disorders^30^; *FCHO1* is involved in clathrin-coat assembly and clathrin-mediated endocytosis and has been implicated in immune deficiency^31,32^. Three novel SHS-PC3 loci were mapped to 62 genes, with five genes in three loci mapped by all four methods. These include *PDE1C* a phosphodiesterase bound by calmodulin that regulates proliferation of vascular smooth muscle cells and may play a pathological role in cardiac remodeling and dysfunction^33^ and *TCF20*, involved in neurodevelopmental diseases and sleep disturbances^34,35^. One novel SHS-PC4 locus was mapped to three genes, including *PCDH17* (mapped by position, eQTL and CI in hippocampal and neural progenitor cells) involved in forming and maintaining neuronal synapses^36^. The two novel SHS-PC5 loci mapped to 71 genes, of which two genes were mapped by all four methods. These include *SLC27A5*, involved in bile acid synthesis and metabolism^37^, which has also been implicated in brain health and neural development^38^. In addition, fifty genes in 28 novel loci have shown drug interactions (Supplementary Table 17). Implicated genes for additional unreported and reported variants are summarized in Supplementary Tables 18 and 19. All significant genes (p<2.64e-6) in gene-based analysis were reported in Supplementary Table 20. Note that genes implicated by fewer mapping methods may also merit prioritization. One intriguing example is that two GWS unreported SHS-PC2-associated variants, rs2821226 (OPTC/ATP2B4; 5.20E-09) and rs1610263 (COL8A1; 4.10E-08) both implicate collagen-pathway genes. Any definitive conclusions based on mapped genes will require functional follow-up.

### Gene set enrichment analyses

We performed pathway enrichment analysis, applying PASCAL^39^ to SHS GWAS summary statistics, and identified significant enrichments of SHS-ADD variants in MAPK and NGF signaling pathways; SHS-PC1 variants in neuronal system, ubiquitin mediated proteolysis, and MAPK signaling pathways; SHS-PC2 in ion transport; SHS-PC3 in circadian, mRNA processing and splicing, G-protein, and metabolic pathways; SHS-PC4 in neuronal system, neurotransmission at synapses, GABA receptor, and long term depression pathways; and SHS-PC5 in the gap junction pathway (Empirical p<5e-4; Fig. 3a and Supplementary Table 21).

**Figure 3.**
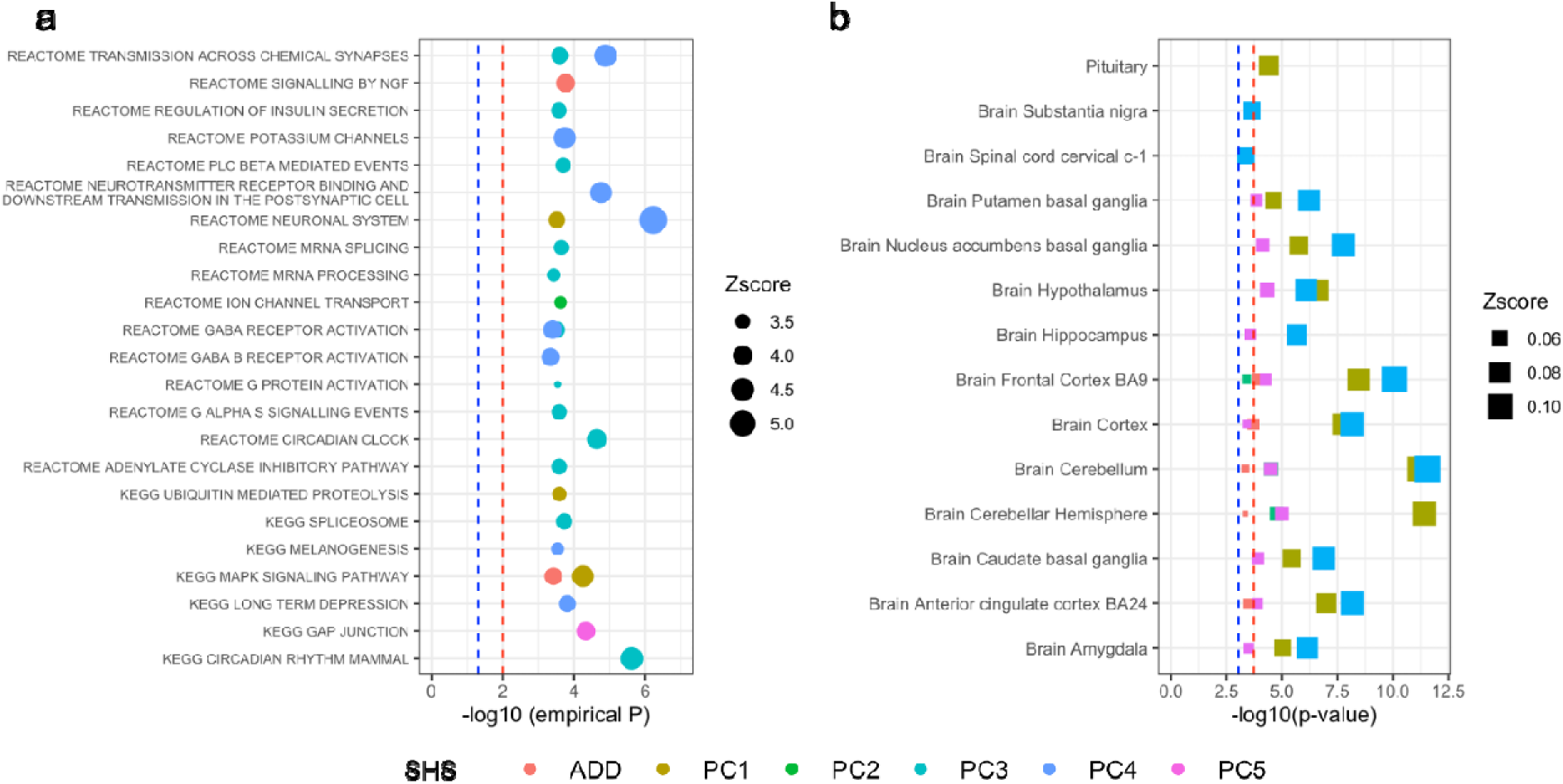
Pathway and tissue enrichment analysis. **a,** Pathway gene sets significantly enriched for SHS genes. **b,** Tissue-specific expression gene sets enriched for SHS genes.

We also performed gene-set enrichment analyses^17^ (tissue, trait, cell type, and pathway) on genes mapped by FUMA using MAGMA (Supplementary Tables 22-24). Tissue enrichment analyses identified multiple brain tissues for all SHS traits except SHS-PC4, with the highest enrichments in cerebellum, hypothalamus, and frontal cortex for SHS-PC1 and frontal cortex, cerebellum, and nucleus accumbens for SHS-PC3. The only non-brain tissue enriched was pituitary tissue for SHS-PC1 (p<0.05/54/6=1.5e-4; Fig. 3b and Supplementary Table 22). Cell type enrichment analysis identified multiple brain cell types for each SHS, and especially for SHS-PC1, including GABAergic neurons in the human midbrain (Supplementary Fig. 9).

Enrichment of SHS-associated genes with phenotype-associated gene sets from the GWAS catalog (Supplementary Tables 23) revealed associations with psychological traits (intelligence, neuroticism, impulsivity/risk-taking, mood, and psychiatric disorders), behavior (e.g., regular activity patterns, alcohol consumption), inflammatory markers and diseases (C-reactive protein, IgG, and inflammatory bowel diseases), blood pressure, adiposity (especially in SHS-PC2 and PC4), and reproductive aging. Gene sets for Alzheimer’s disease and related biomarkers (cerebrospinal fluid tau and amyloid β) were enriched in SHS-PC1, SHS-PC4, and most strongly SHS-PC5. A hippocampal volume gene set was enriched in SHS-PC2 and SHS-PC5. Dendrite gyrus brain volume and kidney disease gene sets were enriched in SHS-PC2. Gene sets associated with intracranial and subcortical brain region volumes, craniofacial microsomia, idiopathic pulmonary fibrosis, and aortic root size were enriched in SHS-PC4. An iron biomarker gene set was enriched in SHS-PC5.

### Genetic and causal relationships between SHS and other common complex traits

LD score regression (LSDC)^40^ revealed numerous phenotypes genetically correlated to SHS traits. Among 375 phenome-wide representative heritable traits (Methods), 256 traits were genetically correlated with at least one SHS (p<0.05/375/6=2.2e-5; Fig. 4 and Supplementary Table 25). Genetic correlations with SHS were strongest (magnitude ∼0.3-0.5) with physical and mental health, and (inversely) with socio-economic status (SES), stress, pain, mental and emotional distress, and recognized health conditions and risk factors. However, these genetic correlations had discernable patterns, unique to each SHS trait. Compared with the SHS-PCs, SHS-ADD had stronger genetic correlations with non-specific health markers, e.g., traits related to overall health, physical conditioning, markers of socio-economic status, healthy lifestyle factors, as well as stronger inverse genetic relationships with pain, activities interfering with sleep, and depression.

**Figure 4.**
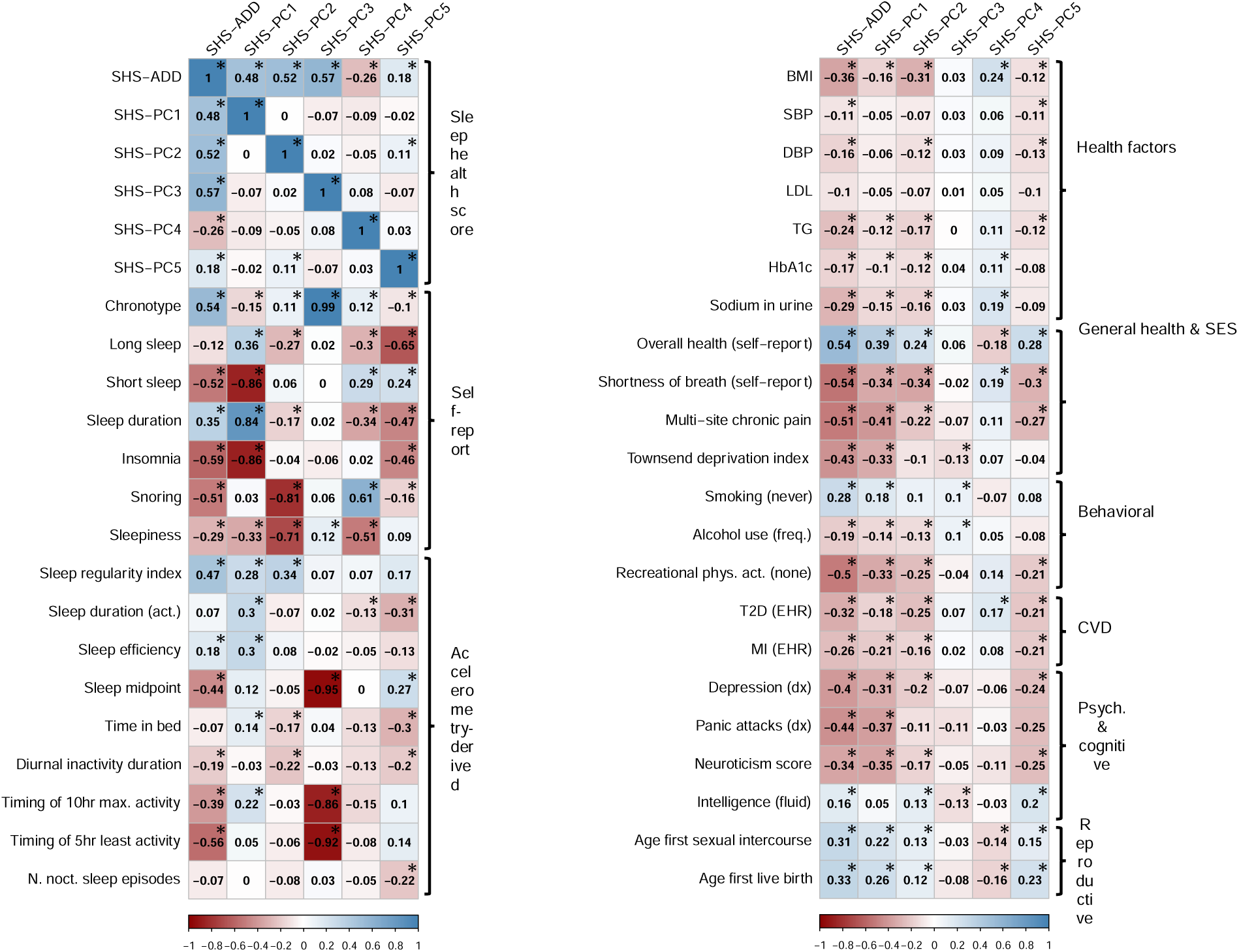
SHS genetic correlations with selected phenotypes. **a,** Sleep traits **(*: p<0.05/132)**. **b,** Selected health outcomes **(*: p<0.05/2250).**

Compared with the other SHSs, SHS-PC1 had stronger inverse genetic correlations with anxiety traits, alcohol addiction, and self-harm behavior; SHS-PC2 had comparatively stronger genetic correlations with daytime napping, diagnosed sleep disorders, and more moderate but, relative to other SHSs, still comparatively stronger inverse genetic correlations with metabolic and adiposity traits. Association patterns for SHS-PC3 differed markedly from other SHS, having lower correlations with overall health and most individual health conditions, inverse association with educational attainment. Compared with the other SHS, genetic correlations involving SHS-PC4 were often inverted, including a (non-significant) inverse association with overall health rating (despite a weak positive phenotypic correlation) as well as positive associations with BMI, and cardiovascular traits, congruent with its positive loading on snoring. SHS-PC5 was preferentially genetically correlated with fluid intelligence, educational attainment, and mother’s age.

We further conducted bidirectional Mendelian randomization (MR) analyses to investigate potentially causal links between sleep health and 50 selected traits (Methods). We identified potential causal effects of lower SHS-PC1 on codeine or tramadol medication use; lower SHS-PC5 on Bipolar disorder; and lower SHS-ADD on smoking initiation and years of schooling (Supplementary Table 26). In the reverse direction, MR identified potential causal effects of greater years of education on higher SHS-ADD and SHS-PC5; lower BMI on higher SHS-PC2; and Alzheimer’s disease risk on higher SHS-PC5 (Supplementary Table 27).

## DISCUSSION

We performed the first large-scale sleep health GWAS investigating five novel PC-based SHSs and compared these with an updated GWAS of SHS-ADD by including related individuals in UKB. Each SHS was based on five underlying self-reported sleep traits: sleep duration, insomnia symptom frequency, daytime sleepiness frequency, chronotype, and snoring, resulting in distinct sleep health composites interpretable via their loadings. The SHS approach emphasizes the co-occurrence of multiple sleep traits, aligning with the multi-dimensional view of sleep health, in which individual components of sleep do not confer sleep health in isolation.

We identified 28 novel significant (p<8.3e-9) loci and 31 additional GWS (p<5e-8) loci that were not reported by previous sleep GWASs. Our findings were supported by sensitivity analysis and PRS associations with clinical sleep outcomes. These loci mapped to genes implicated in neurodevelopment, synaptic signaling, ion channel transportation, cellular energy production, and metabolic processes. The findings collectively suggest that studying SHSs has advanced genetic discovery by linking to plausible biological mechanisms and aligning with established sleep health domains, thereby uncovering novel insights.

Findings for SHS-ADD, combining 5 positive binary sleep traits, suggest a phenotype that captures global sleep health by integrating multiple independent regulatory signals and sleep domains. Notably, SHS-ADD was the SHS most strongly associated with accelerometry-derived sleep regularity index, both phenotypically and genetically, suggesting the conjunction of multiple independent sleep health traits may be a prerequisite for sleep regularity. Genetic correlations were strongest between SHS-ADD and overall health and SES, also correlated strongly with traits related to individual SHS-PCs. SHS-ADD was sensitive to several adjustment factors, including health behaviors and health status, as well as both psychological factors (with SHS-PC1) and BMI (with PC2). Moreover, for SHS-ADD, evidence of enrichments, particularly in neuronal tissues and cell types, was not as strong as for other traits such as SHS-PC1 and SHS-PC3. Together these findings suggest SHS-ADD to be a broad sleep health phenotype, that retains significant heterogeneity, combining multiple independent sleep domains and/or sleep disorders with distinct etiologies. It nevertheless highlights the connections between global sleep health, sleep regularity, and overall health and well-being.

SHS-PC1, longer sleep with less frequent insomnia and sleepiness, reflects correlated self-reported sleep traits across the domains of satisfaction, duration, and alertness, while demonstrating higher heritability than SHS-ADD and each of its underlying sleep traits. SHS-PC1 was the SHS most strongly genetically correlated with accelerometry-based sleep efficiency and duration, suggesting shared genetics with both longer and more efficient objective sleep, with additional genetic correlations suggesting negative relationships with chronic pain, anxiety, and neuroticism. Enrichment analyses identified the ubiquitin proteasome system pathway, important for circadian rhythm regulation and sleep homeostasis^41^, as well as the GABAergic neuronal cell-type, central to neural orchestration of sleep. Overall, these findings suggest a phenotype capturing neurobiological sleep regulation, and support a bidirectional relationship with anxiety^42^ via shared GABAergic regulation^43^, while pointing to further mechanisms involving presence or perception of distressing stimuli, including chronic pain.

SHS-PC2, interpreted as healthy sleep characterized by absent snoring and sleepiness, cardinal symptoms of sleep apnea syndrome, may serve as a surrogate indicating absence of under-diagnosed and poorly captured clinical sleep disordered breathing conditions. Correspondingly, SHS-PC2 showed strong inverse associations, phenotypically and genetically, with adiposity-related measures like BMI, consistent with the strong association between obstructive sleep apnea (OSA) and obesity. Likewise, SHS-PC2 was the SHS most sensitive to BMI adjustment, which nevertheless only modestly attenuated estimated GWAS effects. SHS-PC2 had inverse genetic correlations with cardiometabolic traits, notably type 2 diabetes, which has previously been linked bidirectionally to OSA^44^. Enrichment analyses also link SHS-PC2 with neuronal pathways and hypocampal volume, suggesting neurological involvement in OSA, and with connective-tissue genes (collagen pathway) and traits (adolescent idiopathic scoliosis, aortic root size), consistent with a role for connective tissue in pharyngeal collapsibility related to sleep-disordered breathing^45^.

SHS-PC3 largely recapitulates chronotype and confirmed known associations with circadian genes and pathways. It is notable that chronotype was largely independent not only from other SHS-PCs but was also more weakly genetically correlated with phenome-wide health outcomes, while being moderately genetically correlated with healthy lifestyle behaviors, such as physical activity and time spent outdoors in summer.

Though SHS-PC4 and SHS-PC5 have more complex interpretations due to both positive and negative loadings on healthy sleep traits, they were nonetheless roughly as heritable as the least heritable individual trait (sleepiness) and contributed novel genetic findings. SHS-PC4, interpreted as snoring without sleepiness, showed genetic enrichment in neurotransmission pathways and craniofacial structure, suggesting mechanisms that could lead to sleepiness without snoring, and/or snoring without sleepiness. The latter would be consistent with a sleep-disordered breathing phenotype resulting from reduced craniofacial dimensions that cause pharyngeal narrowing and turbulent airflow (or snoring) without the severe airway collapsibility, sleep fragmentation, and inflammation characteristic of obstructive sleep apnea syndrome ^46,47^.

SHS-PC5, characterized by short sleep without insomnia, was genetically correlated with later objective sleep midpoint and fewer accelerometry-measured sleep episodes, as well as positive genetic correlations with markers of cognitive decline, including memory loss, cerebrospinal fluid t-tau levels, and Alzheimer’s disease, and with enrichment of genes in the gap junction pathway, implicated in amyloid-β clearance by astrocytes in Alzheimer’s disease^48^. This suggests that SHS-PC5 may characterize shorter, delayed sleep, without insomnia symptoms, indicative of accelerated brain aging, rather than a natural short sleep or ‘super-sleeper’ phenotype^3^. The low likelihood of insomnia in this subtype is consistent with lack of consistent data implicating insomnia in cognitive decline, potentially due to the heterogeneity of conditions underlying insomnia.

Colocalization analyses revealed selective cases of shared regulation, but also pointed to sometimes differing relationships with underlying components, particularly in the special cases of PC2 vs PC4, and PC1 vs PC5, which share underlying traits with opposite loadings. For example, *DLEU7*, proximal to rs592333, colocalized opposite associations with PC2 and PC4, consistent with a role in snoring (which loaded positively in PC4 and negatively in PC2) and consistent with prior associations of the *DLEU7* locus with adiposity^49^. Conversely, rs1846644 colocalized positive associations with both PC2 and PC4 at the *KSR2* locus, a gene highly expressed in cerebellar Purkinje neurons^50^, suggesting a link to sleepiness regardless of the presence of snoring, in keeping with a role for Purkinje neurons in sleep-wake transition^51^. Similarly, for SHS-PC1 and PC5, *PAX8* colocalized with opposite directions, in keeping with opposite loadings on sleep duration, while *MEIS1* colocalized with similar direction in keeping with consistent loading on insomnia.

Several additional findings pointed to mechanisms shared across SHS indicating broad-based involvement in sleep health. For example, PC2 and PC4 at the *KSR2* locus, a gene highly expressed in cerebellar Purkinje neurons, which were again implicated in PC1 by two novel loci, *ANKFY1* and *WDR73/ZNF592*, suggesting cerebellar regulation of sleep maintenance efficiency. Additional findings consistently reinforced the role of neural development, as well as consistently implicating neurotransmitters and synaptic signaling. The association of SHS-PC2 with *FCHO1* at rs12979056 gives further support for a role of clathrin-coat vesicle transport in sleep health, presumably in synaptic function, a mechanism previously implicated by the

*STON1-GTF2A1L, TOR1A, TOR1B, AP2B1* (PC3) and *AP3B2* (PC1) loci. Pathways and genes related to MAPK, GAP-junction, immune signaling, and energy metabolism processes were found to associate across multiple SHS. Interestingly, the two identified novel functional variants, rs8074498 in PC1 and rs138572890 in PC5, the PC phenotypes loading on duration/insomnia, were connected to glucose transporters (respectively *GLUT4* and *GLUT1*). Moreover, enrichment analysis for individual SHS all highlighted gene expression in brain tissues and cells, and associations with metabolic, inflammatory, and psychiatric traits, which reinforce critical roles for central nervous system, metabolic, and immune system function on sleep regulation. Multi-trait genetic correlation analyses further suggested more broadly interrelatedness of sleep health with overall health, lifestyle, behavioral and psychological traits, as well as pain, physical frailty, and deconditioning. However, some caution is warranted as some apparent genetic relationships could be induced by factors relating to the subjective rating of sleep health, remaining heterogeneity within SHS traits, or unadjusted confounders, including remaining population stratification^52^.

This study has several limitations. First, SHS-PCs incorporated sleep traits in a linear fashion, which may not reflect the complexity of their contribution to sleep health, e.g., the U-shaped contribution of sleep duration as modeled in SHS-ADD. Second, SHS-PCs only focus on five self-reported sleep questions to maximize sample size and to be more comparable to SHS-ADD as previously published on in UKB by using the same underlying sleep phenotypes. This could limit our ability to fully capture a comprehensive sleep health composite, due to limitations of subjective assessment or the ability of these questions to fully capture well being in sleep health. Third, the data-driven PC approach may limit generalizability across different studies and populations. We were able to identify HCHS/SOL as having similar questionnaire data, however, generalization to this cohort was challenging due to population and age differences.

Correspondingly it was not surprising that PC analysis performed in this cohort resulted in different loadings. A potentially productive approach to future validation studies would be to average PC loadings across different studies in a meta-analysis^53^. Lastly, we note that the SHS-PCs were based on phenotypic correlations, which could limit the ability to derive composite phenotypes of maximum heritability. However, in a sensitivity analysis (data not shown) we constructed sleep scores informed by genetic correlations using the ‘maxH’ maximally heritable approach^54^, for which the loadings and heritability of the derived phenotypes were numerically and qualitatively similar. Nonetheless, genetic correlation information could be valuable in future research integrating additional objective sleep-related phenotypes.

In summary, this study introduces a novel approach to understanding sleep genetics using PC-based SHS, which effectively distinguishes differing mechanisms of distinct, domain-specific sleep-related traits. In keeping with the large influence of insomnia (related to SHS-PC1) and sleep apnea (related to PC2) on sleep health in the population, along with the independent role of chronotype (PC3), our approach appears to have enhanced genetic discovery by separately targeting these domain-specific sleep health scales. Future research involving SHSs built from objective data may provide enhanced targeting of psychosocial domains and neuroregulatory sleep mechanisms, resulting in further genetic discovery.

## METHODS

### Population and study design

The discovery analysis was conducted on participants of European ancestry from the UK Biobank study^55^. The UK Biobank is a prospective study that has enrolled over 500,000 people aged 40-69 living in the United Kingdom. Baseline measures collected between 2006 – 2010, including self-reported heath questionnaire and anthropometric assessments, were used in this analysis. Participants taking any self-reported sleep medication (described before^7,8,10^) were excluded. The UK Biobank study was approved by the National Health Service National Research Ethics Service (ref. 11/NW/0382), and all participants provided written informed consent to participate in the UK Biobank study.

### Genotype

DNA samples of 502,631 participants in the UK Biobank were genotyped on two arrays: UK BiLEVE (807,411 markers) and UKB Axiom (825,927 markers). 488,377 samples and 805,426 genotyped markers passed standard QC^56^ and were available in the full data release. 452,071 individuals of European ancestry (based on K-means clusters on genomics PCs) were studied with available phenotypes and genotyping passing quality control. SNPs were imputed to a combined Haplotype Reference Consortium (HRC) and 1000 Genome panel. SNPs with minor allele frequency (MAF)>0.001, BGEN imputation score >0.3, maximum per SNP missingness of 10%, and samples with a per-sample missingness of 40% were kept in the GWAS.

### Sleep trait assessment

The UK Biobank baseline questionnaire assessed chronotype, sleep duration, insomnia symptoms, snoring, and excessive daytime sleepiness via self-report responses. Self-reported sleep duration was recorded as an integer-valued variable via responses to the question “About how many hours sleep do you get in every 24-LJhours? (please include naps).” The remaining questions had ordinal responses, which were assigned to an integer scale as follows. Chronotype (morningness): “Do you consider yourself to be:” -2. Definitely an ’evening’ person; -1. More an ’evening’ than a ’morning’ person; 0. Do not know; 1. More a ’morning’ than an ’evening’ person; 2. Definitely a ’morning’ person; NA. Prefer not to answer. Insomnia Symptoms: “Do you have trouble falling asleep at night, or wake up in the middle of the night?” 1. Never/rarely; 2. Sometimes; 3. Usually; NA. Prefer not to answer. Snoring: “Does your partner or a close relative or friend complain about your snoring?” 1. Yes; 0. No; NA. Do not know or Prefer not to answer. Subjective daytime sleepiness: “How likely are you to doze off or fall asleep during the daytime when you don’t mean to? (e.g. when working, reading, or driving?)” 0. Never/rarely; 1. Sometimes; 2. Often; 3. All of the time; NA. Do not know or Prefer not to answer. Individuals with any missingness (NA) from any sleep questionnaires were excluded from the analysis.

### Sleep health score construction

For the UK Biobank additive sleep health score (SHS-ADD), consistent with previous studies^13,14^ we assigned one point to each of five positive sleep traits, as follows: Chronotype: More a ’morning’ than an ’evening’ person or Definitely a ’morning’ person; Sleep Duration: from 7 to 8 hours (inclusive); Insomnia Symptoms: Never/rarely; Snoring: No; Subjective daytime sleepiness: 0. Never/rarely. No subjects were excluded; those who did not report the positive attributes were coded as zero for that trait. The final SHS-ADD rating is the total number of positive sleep traits for each individual on a scale of 0-5. We performed principal component analysis of the above-described integer-scale self-report sleep question responses, after centering and scaling to standardize each response to mean zero, variance one, to compute SHS-PC1 through SHS-PC5. SHS-PCs were oriented so that they were positively correlated with self-assessed overall health (UKB Data-Field 2178: Overall health rating).

### Covariate measurements

Covariates used in the sensitivity analyses included potential confounders (depression, socio-economic deprivation based on residential area, alcohol intake frequency, smoking status, caffeine intake, employment status, marital status, neurodegenerative disorders, and use of psychiatric medications). Depression was recorded as a binary variable (yes/no) corresponding to question “Ever depressed for a whole week?”. Social economic status was measured by the Townsend Deprivation Index based on aggregated data from national census output areas in the UK. Alcohol intake frequency was coded as a continuous variable corresponding to “daily or almost daily”, “three or four times a week”, “once or twice a week”, “once to three times a month”, “special occasions only”, and “never” drinking alcohol. Smoking status was categorized as “current’, “past”, or “never” smoked. Caffeine intake was coded continuously corresponding to self-reported cups of tea/coffee per day. Employment status was categorized as “employed”, “retired”, “looking after home and/or family”, “unable to work because of sickness or disability”, “unemployed”, “doing unpaid or voluntary work”, or “full or part-time student”. Neurodegenerative disorder cases (N=517) were identified as a union of International Classification of Diseases (ICD)-10 coded Parkinson’s disease (G20-G21), Alzheimer’s disease (G30), and other degenerative diseases of nervous system (G23, G31-G32).

### Accelerometry data

Accelerometry data were collected using Axivity AX3 wrist-worn triaxial accelerometers in 103,711 individuals from the UK Biobank for up to 7 days, 3-10 years after baseline^57^. Sleep period time (SPT)-window and activity levels were extracted using a heuristic algorithm using the R package GGIR (https://cran.r-project.org/web/packages/GGIR/GGIR.pdf)^58^. Briefly, for each individual, a 5-minute rolling median of the absolute change in z-angle (representing the dorsal-ventral direction when the wrist is in the anatomical position) across a 24-hour period. The 10th percentile of the output was used to construct an individual’s threshold, distinguishing periods with movement from non-movement. Inactivity bouts were defined as inactivity of at least 30 minutes duration. Inactivity bouts with gaps of less than 60 minutes were combined into blocks. The SPT-window was defined as the longest inactivity block, with sleep onset as the start of the block and waking time as the end of the block. This algorithm provides comparable estimates of sleep onset time, waking time, SPT-window duration, and sleep duration within the SPT-window with polysomnography derived metrics^58^. After quality control based on missingness, wear time, and calibration, eight metrics were generated and analyzed in this study, namely: M10 (midpoint of the 10 consecutive hours of maximum activity), L5 (midpoint of the 5 consecutive hours of minimum activity), sleep midpoint, sleep duration, sleep efficiency, diurnal inactivity, number of nocturnal sleep episodes, and sleep regularity index (accounting for wake after sleep onset and daytime napping^59^).

### Genome-wide association analysis

We performed a genome-wide association analysis (GWAS) of six SHS traits as continuous variables using linear mixed regression models adjusting for age, sex, genotyping array, 10 PCs, and genetic relatedness matrix in BOLT-LMM. Reference 1000 genome European-ancestry (EUR) LD scores and genetic map (hg19) were utilized in this analysis. X-chromosome data were imputed and analyzed separately (with males coded as 0/2 and female genotypes coded as 0/1/2) using the same analytical approach in BOLT-LMM as was done for analysis of autosomes. FUMA was used to annotate the genome-wide significant (GWS) risk loci (p<5e-8), lead independent SNPs (r^2^<0.1), and all SNPs in LD with independent SNPs (r^2^>=0.6) within a genomic region (including ANNOVAR functional consequence, CADD score, RegulomeDB score, as well as 15 chromatin states, and GWAS Catalog associations). We compared the GWS loci to loci reported by biobank-based GWAS of sleep traits published by June 2022, including insomnia^8,9^, sleep duration^7^, daytime sleepiness^10^, chronotype^12^, snoring^11^, daytime napping^60^, obstructive sleep apnea (OSA)^61^, restless legs syndrome (RLS)^62^, and prior SHS-ADD in UKB unrelated individuals^13^. GWS loci at least 500kb from any of the reported loci were noted as unreported. To account for performing six simultaneous GWAS, we report significant novel loci defined as the unreported loci passing a stricter significant threshold correcting for six traits (p<8.3e-9).

The constructed SHS traits, as sums of ordinal and integer-valued variables, were somewhat non-normal, which has the potential to affect Type-I error. Consistent with prior UKB-GWAS of ordinal sleep phenotypes, conducting GWAS using linear mixed models implemented in BOLT is expected to mostly ameliorate this concern. BOLT has previously been shown to preserve Type-1 error adequately for non-normal ordinal and binary phenotypes, as long as the sample sizes in different groups not markedly imbalanced.^63^

### Gene mapping and gene-based analysis

We used FUMA to map SNPs to genes using three methods: positional mapping (<=10kb), cis-eQTL (<=1Mb) in GTEx v8 tissues (FDR<0.05), and 3D Chromatin interaction (CI) in 127 tissue/cell types (FDR<1e-6). We also looked up ensemble phenotypes using R biomaRt package and drug interaction evidence using DGIdb for mapped genes. We next performed gene-based association analysis using genome-wide summary statistics using MAGMA^17^ in FUMA. Input SNPs were mapped to 18,931 protein coding genes. Genome-wide significance level was defined as p<0.05/18,931=2.641e-6.

### Gene set enrichment analysis

We performed pathway enrichment analysis using PASCAL, which estimated a combined association p-value from the summary statistics of multiple SNPs in a gene^39^. Significant KEGG, Reactome, and BIOCARTA pathways for each SHS trait were identified using empirical p<0.05. We also performed gene set enrichment analysis on positional, eQTL and CI mapped genes in FUMA MAGMA^17^ adjusted for gene size. Significant tissues were identified using p<0.05/54/6 accounting for 54 tissues in GTEx v8 and six traits. Significant pathways (KEGG, Reactome, and GO pathways in MSigDB), and GWAS gene set (GWAS Catalog) were identified using adjusted p<0.05. PASCAL pathway enrichment incorporates effect sizes and LD at individual loci, whereas FUMA MAGMA gene set enrichment is based only on the overlap in sets of associated genes and does not account for LD, impacting interpretation.

### Colocalization analysis

We performed colocalization analysis across SHS traits using HyPrColoc^64^ and genome-wide summary statistics to assess the shared genetic risk factors. We expected that SHS-PCs had less colocalized loci given their independency. Colocalized loci among SHS-PCs should have true pleiotropic effect and play a central regulation role.

### Sensitivity analyses

Sensitivity analyses of all GWS loci were performed additionally adjusting for potential confounders (including adiposity, socio-economic status, alcohol intake frequency, smoking status, caffeine intake, employment status, marital status, and psychiatric problems) individually in 308,902 unrelated individuals using PLINK in additional to adjusting for age, sex, genotyping array and 10 PCs in PLINK 1.9. We used a hard-call genotype threshold of 0.1, SNP imputation quality threshold of 0.80, and a MAF threshold of 0.001. We also performed the analysis excluding shift workers and individuals with chronic health or psychiatric illnesses (N=115,297) and in males (N=145,196) and females (N=163,716) (without adjusting for sex).

### Genetic risk score analysis

We constructed a weighted GRS comprised of GWS loci for each SHS and tested for associations with other self-reported sleep traits (sleep duration, long sleep duration, short sleep duration, insomnia, chronotype, and snoring), and 7-day accelerometry traits in the UK Biobank. Weighted GRS analyses were performed by summing the products or risk allele count multiplied by the effect estimate reported in the SHS GWASs using R package gds (https://cran.r-project.org/web/packages/gds/gds.pdf). We also tested the GRSs of reported loci for sleep traits using the same approach.

Both individual loci and PRS associated with SHS tended to associate with multiple self-reported sleep traits from which they were derived, as well as objective accelerometry-derived sleep traits, which were not used in SHS construction. For example, the *MEIS1* locus (SHS-ADD, SHS-PCs 1, 3, and 5) was associated with accelerometry-derived circadian/sleep timing, sleep duration, efficiency, and regularity, and the *PAX8* (SHS-PCs 1, 2, and 5) locus was associated with accelerometry sleep duration, efficiency, and daytime inactivity (p<5e-8; Supplementary Table 9). Similarly, GRSs for SHS-ADD and SHS-PC1 associated with longer accelerometry-derived sleep duration, and higher sleep efficiency and regularity, while GRS of SHS-PC2 associated with shorter accelerometry-assessed daytime inactivity (p<0.05/15/6=5.6e-4; Supplementary Table 10).

### Genetic correlation analysis

We estimated genetic correlations among SHS and with other self-reported and accelerometry sleep traits using LDSC^40^ and genome-wide SNPs mapped to the HapMap3 reference panel. To understand the genetic overlap with a range of common health problems, we selected 381 representative UKB traits by choosing the 232 most heritable traits using UKB hierarchical phenotype categories (selecting at most one ‘level’ per phenotype, at most 5 phenotypes per phenotype category, and at most 25 phenotypes per category group), as well as all 195 (partly overlapping) traits in the PanUKBB maximally independent set of phenotypes (https://pan.ukbb.broadinstitute.org/blog/2022/04/11/h2-qc-updated-sumstats/index.html). Of the 381 selected traits, 375 passed heritability thresholds and were carried on for LDSC analyses. Traits passed heritability QC if stratified LD score regression in the UKB Europeans defined in the Pan UKBB GWAS (sldsc_25bin_h2_pval_EUR) was significant with p<0.05/381. Multiple-testing-corrected significance level for genetic correlation was defined as p<0.05/375/6=2.2e-5.

### Mendelian Randomization analysis

Bidirectional mendelian randomization (MR) analyses, as implemented in the *TwoSampleMR* R package^65^, were conducted to investigate potentially causal links between sleep health and 50 representative traits from across the phenome, selected (prior to performing MR) based on their relationships with sleep traits, either in prior literature, or based on results of interest from the genetic correlation analysis. The final list of traits (below) was also determined by manual review of availability of non-UKB GWAS summary statistics in the IEU open GWAS project database. We report associations that were significant correcting for testing 50 phenotypes and 6 sleep health traits (p<0.05/300), under 2-sample inverse variance weighted (IVW) methodology, while also requiring effects estimated under MR-Egger to be consistent in direction, and that the putative causal direction was not invalidated in a Steiger directionality test, such that the selected instruments for the exposure had a greater R^2^ variance explained in the exposure phenotype than the outcome phenotype (neither the MR-Egger nor Steiger tests were required to be statistically significant). SNP heterogeneity, for which horizontal pleiotropy is a likely cause, was evaluated by Cochran’s Q (for IVW)^66^ and Rücker’s Q’ (for MR-Egger)^67^ statistics. Horizontal pleiotropy was assessed by MR-Egger intercept.

MR traits: finn-b-R18_COUGH: Cough, finn-b-RX_CODEINE_TRAMADOL: Codeine or tramadol medication, ieu-a-73: Waist-to-hip ratio, ieu-b-103: HbA1C, finn-b-F5_NEUROTIC: Neurotic, stress-related and somatoform disorders, ieu-a-832: Rheumatoid arthritis, ieu-a-962: Ever vs never smoked, finn-b-G6_PARKINSON_EXMORE: Parkinson’s disease (more controls excluded), ieu-a-44: Asthma, ieu-b-18: multiple sclerosis, ieu-a-89: Height, finn-b-KRA_PSY_ANXIETY: Anxiety disorders, ebi-a-GCST002216: Triglycerides, finn-b-R18_PAIN_THROAT_CHEST: Pain in throat and chest, ieu-b-2: Alzheimer’s disease, finn-b-ANTIDEPRESSANTS: Depression medications, finn-b-Z21_PERSONS_W_POTEN_HEALTH_HAZARDS_RELATED_SOCIO_PSYCHOSO_CIRCUMSTANC: Persons with potential health hazards related to socioeconomic and psychosocial circumstances, finn-b-K11_DIVERTIC: Diverticular disease of intestine, ieu-a-113: Neo-agreeableness, ieu-a-115: Neo-extraversion, ieu-a-1009: Subjective well-being, finn-b-PAIN: Pain (limb, back, neck, head abdominally), finn-b-ALCOHOL_RELATED: Alcohol related diseases and deaths, all endpoints, ieu-b-4855: FEV1/FVC, ieu-a-294: Inflammatory bowel disease, ieu-a-16: Childhood intelligence, finn-b-F5_SUBSNOALCO: Substance use, excluding alcohol, finn-b-F5_PANIC: Panic disorder, ieu-a-114: Neo-conscientiousness, ieu-b-4859: Physical activity, ieu-a-116: Neo-neuroticism, finn-b-F5_GAD: Generalized anxiety disorder, finn-b-I9_IHD: Ischaemic heart disease, wide definition, finn-b-F5_DEPRESSIO: Depression, ieu-b-4820: Age at first birth, ebi-a-GCST002223: HDL cholesterol, ieu-a-1001: Years of schooling, finn-b-RX_PARACETAMOL_NSAID: Paracetamol of NSAID medication, ebi-a-GCST003116: Coronary artery disease, ieu-b-38: systolic blood pressure, ieu-b-41: bipolar disorder, finn-b-M13_ENTESOPATHYLOW: Enthesopathies of lower limb, excluding foot, ieu-b-73: Alcoholic drinks per week, finn-b-K11_REFLUX: Gastro-oesophageal reflux disease, finn-b-E4_DIABETES: Diabetes mellitus, ieu-a-1095: Age at menarche, ieu-a-835: Body mass index, ieu-a-117: Neo-openness to experience, ieu-b-39: diastolic blood pressure, ebi-a-GCST002222: LDL cholesterol

### Replication analysis

HCHS/SOL is a community-based study in the U.S., which includes 16,415 adults aged 18-74 with self-identified Hispanic/Latino background^68,69^. Individuals were recruited from randomly selected households near four centers in Miami, San Diego, Chicago and the Bronx area of New York. Self-reported sleep duration, insomnia (assessed by Women’s Health Initiative Insomnia Rating Scale [WHIIRS]), daytime sleepiness (assessed by Epworth Sleepiness Scale [ESS]), and snoring were collected in 13,268 individuals at baseline. Chronotype was only available in 1,855 individuals enrolled in the Sueño sleep ancillary study.

We converted the sleep data in HCHS/SOL to UKB scale. For insomnia, we used two questions that were part of the WHIIRS questionnaire in the HCHS/SOL: 1) “Did you have trouble falling asleep?” 2) “Did you wake up several times at night?”. Each question provided 5 choices: 1. No, not in the past four weeks; 2. Yes, less than once a week; 3. Yes, 1 or 2 times a week; 4. Yes, 3 or 4 times a week; 5. Yes, 5 or more times a week. We converted the sum score of the two questions (2-10) to UKB scale as: 2-4=“Never/rarely”; 5-8=“Sometimes”; 9-10=“Usually”. For sleepiness, we converted the ESS score (0-24) to UKB scale as: 0-10=“Never/Rarely”; 11-14=“Sometimes”; 15-18=“Often”; 19-24=“All the time”. For snoring, we converted the 4-level answers for the question “How often do you snore now?” to UKB scale as: 0 if the answer was “Never” or “Rarely” and 1 if the answer was “Sometimes” or “Always”. Chronotype was collected in Sueño using the same questionnaire to UKB. We performed multiple imputation to calculate and impute the missing chronotype in the rest of the samples in HCHS/SOL (N=11,413) using chained equations method with linear regression on relevant variables, specifically, sex, age, BMI, and four sleep timing questions “What time do you usually go to bed in the weekday?”, “What time do you usually go to bed in the weekend?”, “What time do you usually wake up in the weekday?”, and “What time do you usually wake up in the weekend?”. We found the post-imputation distribution of the chronotype responses matched those observed in Sueño. We then constructed SHS in HCHS/SOL using the same loadings from UKB.

Of the 13,268 individuals with imputed phenotype data, 11,144 individuals with genotype data and consented to genetic research are available for replication. Genotyping was conducted using an Illumina Omni2.5M SNP array with additional customized content, including 2,536,661 SNPs and imputed to TOPMed reference panel using TOPMed imputation server. Genetic association analysis for the 400 GWS loci were performed using linear mixed model in R Genesis software adjusting for age, sex, study center, sampling weights, five principal components of genetics representing ancestry, with random effects corresponding to kinship, household, and block unit.

### Validation of SHS PRS on clinical phenotypes in a clinical biobank

The Mass General Brigham (MGB) Biobank is a clinical biobank enriched for disease states supplemented with genetic data from the MGB healthcare network in Massachusetts. Since 2009, patients have been recruited through online channels or in person from various MGB community-based primary care facilities and specialty tertiary care centers. Among the enrolled patients, a subset (n = 64,639) provided blood samples for genotyping. DNA extracted from samples was genotyped using the Infinium Global Screening Array-24 version 2.0 (Illumina).

Imputation was carried out through the Michigan Imputation server with the Trans-Omics for Precision Medicine (TOPMed) (version r2) reference panel, and haplotype phasing was performed using Eagle version 2.3. Low-quality genetic markers and samples were excluded^70^. Pairs of related individuals (kinship > 0.0625) were identified, and one sample from each related pair was excluded^70^. To correct for the population substructure, principal components of ancestry were computed using TRACE and the Human Genome Diversity Project^71,72^. Individuals with non-European ancestry were excluded to limit genetic heterogeneity in the present analysis.

Among 47,082 adult patients included in the present analysis (mean age = 60.4 ± 17.0; 53.8% female), 6 polygenic risk scores for the SHS PCs and additive model were generated using Polygenic Risk Score–Continuous Shrinkage^73^. Each score was standardized with a mean of 0 and a standard deviation (SD) of 1. Case ascertainment for sleep disorders were based on clinical phenotypes identified from ICD-9/-10 billing codes and mapped to PheWAS codes (i.e., “phecodes”) based on clinical similarity generated by the PheWAS R package^74^. A total of 13 sleep disorders were considered in the analysis. For each disorder, participants with at least two codes were set as cases, and those with no relevant codes were set as controls. Associations between the PRSs and each disorder were tested using logistic regressions adjusted for age, sex, genotyping array, batch, and PCs of ancestry. Significance was determined using Bonferroni-adjusted P values for the total number of tests (0.05/(6 PRSs x 13 phenotypes) = 6.4e-4).

## Data Availability

All data produced in the present study are available upon reasonable request to the authors.

## Acknowledgements

This research has been conducted using the UK Biobank Resource under application 6818. We would like to thank the participants and researchers from the UK Biobank who contributed or collected data. This work was supported by the National Institute of Health (NIH) grants R01HL153814 (to H.W.), R01HL146751 (to R.S.), R35HL135818 (to S.R.), and NIHR Manchester Biomedical Research Centre. S.D.K. is supported by the National Institute of Health and Care Research (NIHR) Oxford Health Biomedical Research Centre, NIHR Efficacy and Mechanisms Evaluation Programme (Ref: 131789), NIHR Programme Grants for Applied Research (Ref: 203667) and the Wellcome Trust (226784/Z/22/Z and 227093/Z/23/Z). D.A.L. is supported by the UK Medical Research Council (MC_UU_00032/05), British Heart Foundation (CH/F/20/90003 and AA/18/1/34219) and Diabetes UK (17/0005700). J.I.R. is supported by the National Center for Advancing Translational Sciences, CTSI grant UL1TR001881, and the National Institute of Diabetes and Digestive and Kidney Disease Diabetes Research Center (DRC) grant DK063491 to the Southern California Diabetes Endocrinology Research Center. The Hispanic Community Health Study/Study of Latinos was supported by contracts from the National Heart Lung and Blood Institute to the University of North Carolina (N01-HC65233), University of Miami (N01-HC65234), Albert Einstein College of Medicine (N01-HC65235), the University of Illinois at Chicago (HHSN268201300003I), Northwestern University (N01-HC65236), and San Diego State University (N01-HC65237). The following Institutes/Centers/Offices contributed to the HCHS/SOL through a transfer of funds to the National Heart Lung and Blood Institute: National Center on Minority Health and Health Disparities, the National Institute of Deafness and Other Communications Disorders, the National Institute of Dental and Craniofacial Research, the National Institute of Diabetes and Digestive and Kidney Diseases, the National Institute of Neurologic Disorders and Stroke, and the Office of Dietary Supplements.

