## Supplementary Fig for "Genome-wide association analysis of composite sleep health scores in 413,904 individuals"

Supplementary Figures

### Supplementary Fig. 1. Histograms of SHS traits.

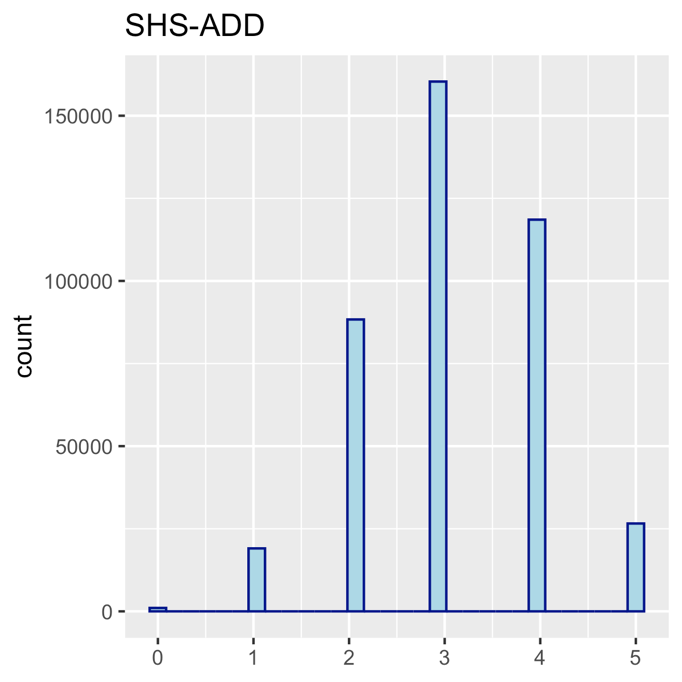

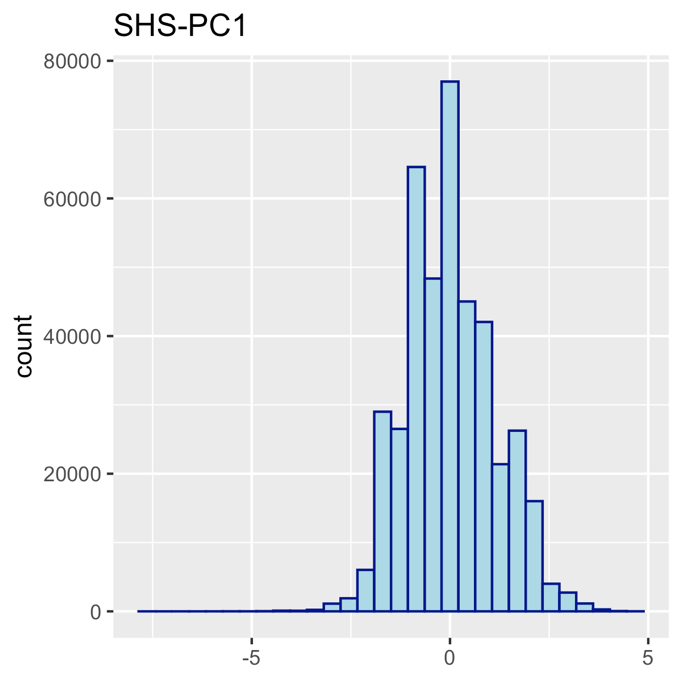

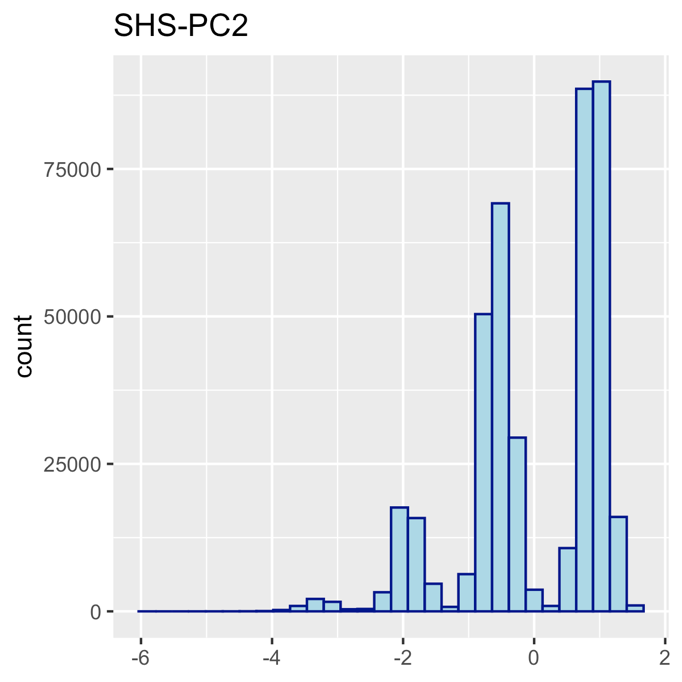

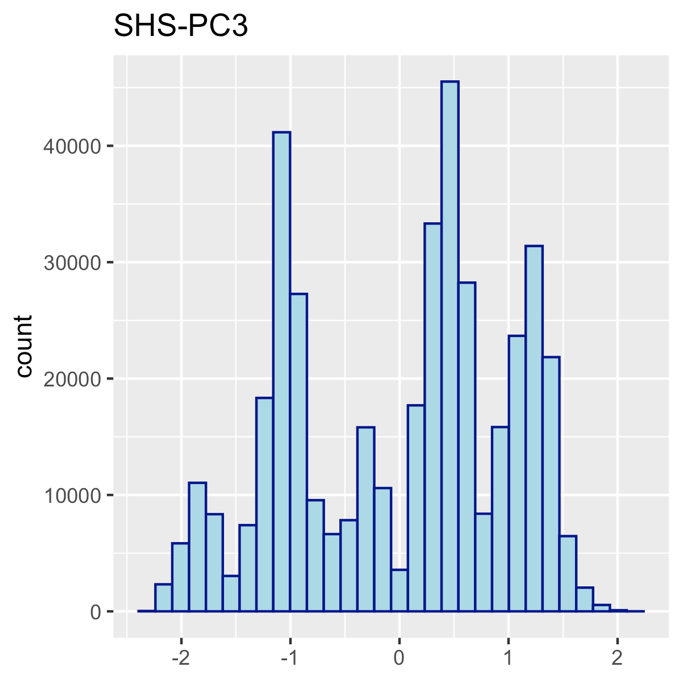

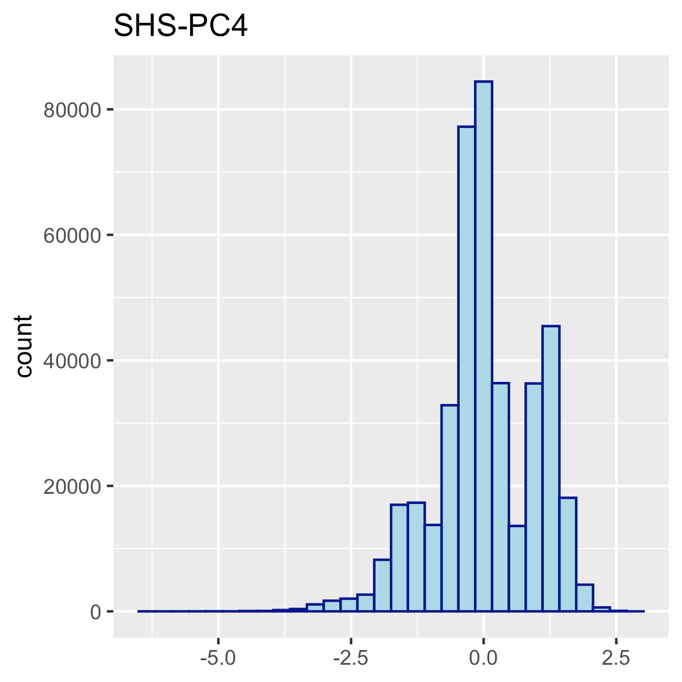

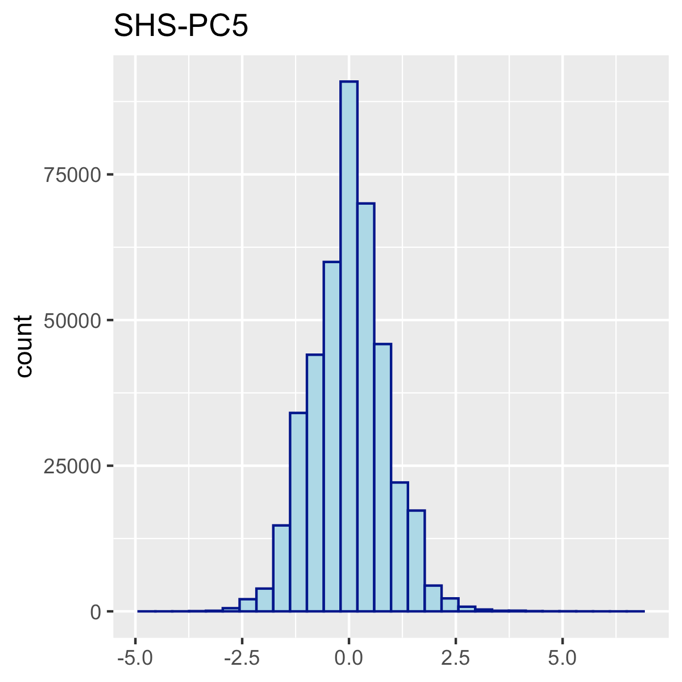

### Supplementary Fig. 2. Phenotypic and genetic correlations between SHS traits with self-reported and accelerometry sleep traits in UKB.

a. Spearman correlations with self-reported and accelerometry sleep traits (*: p<3.8e-04).

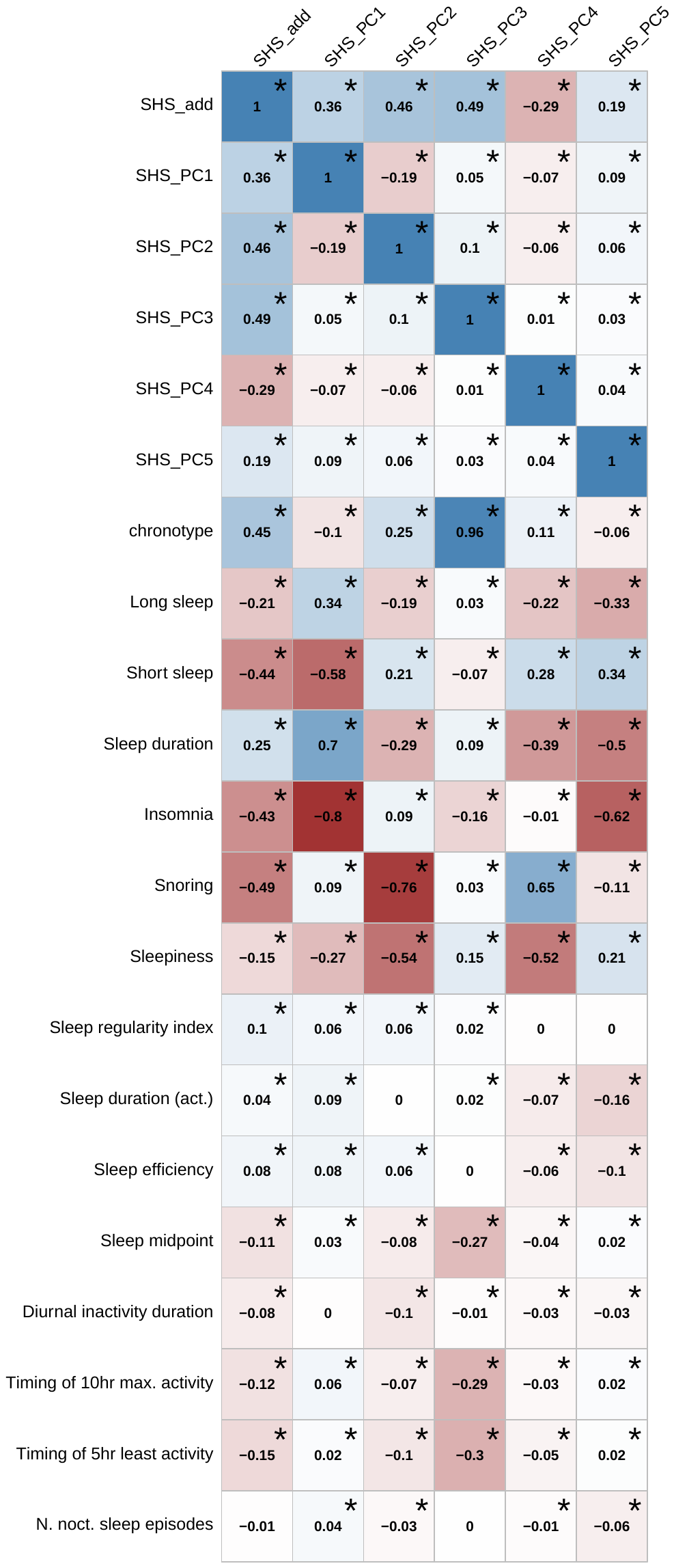

b. LDSC genetic correlations with self-reported and accelerometry sleep traits (*: p<3.8e-04).

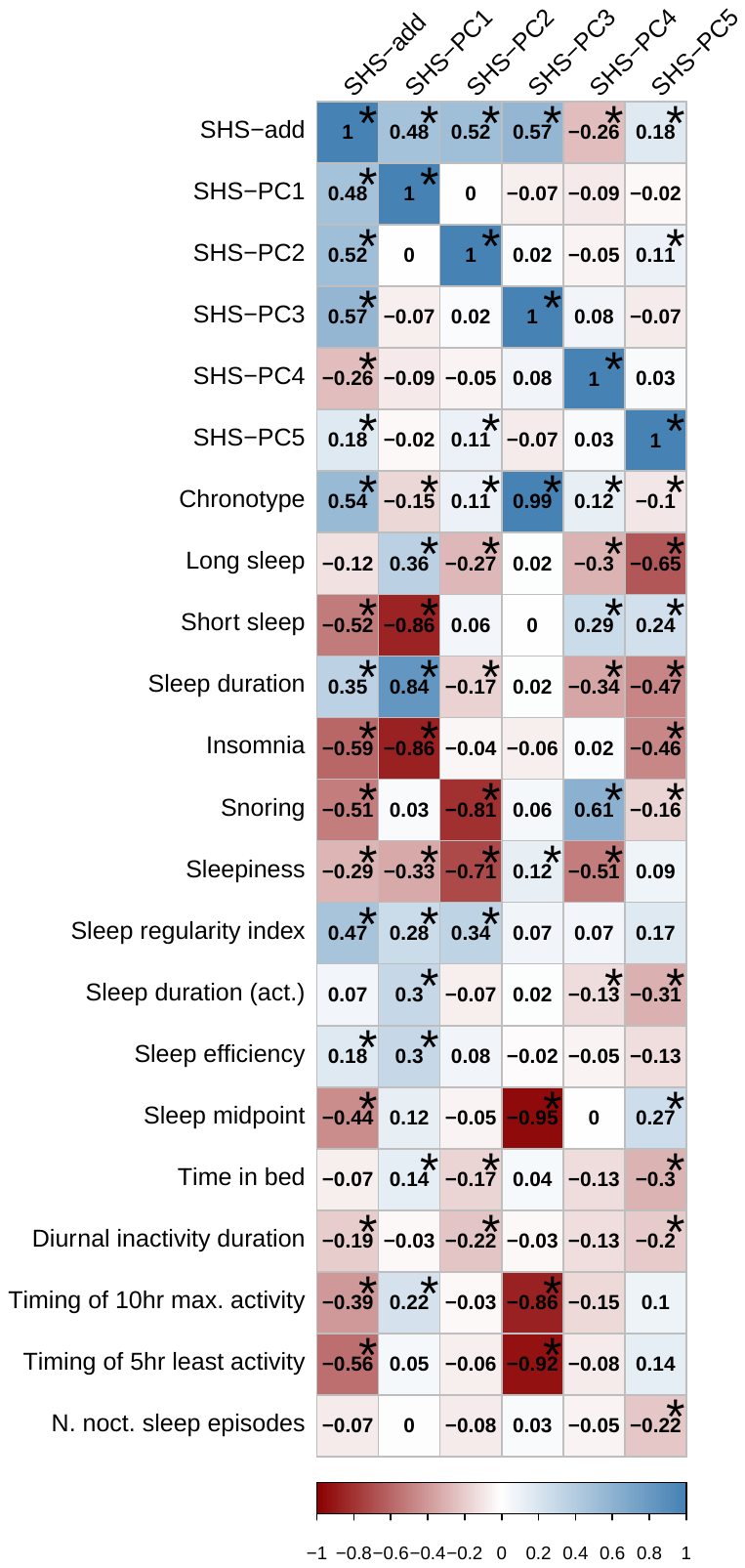

### Supplementary Fig. 3. Phenotypic and genetic correlation plots of SHS traits with selected representative traits

a. Spearman rank correlations of SHSs with phenotypic traits of interest (*: p<2.2e-05).

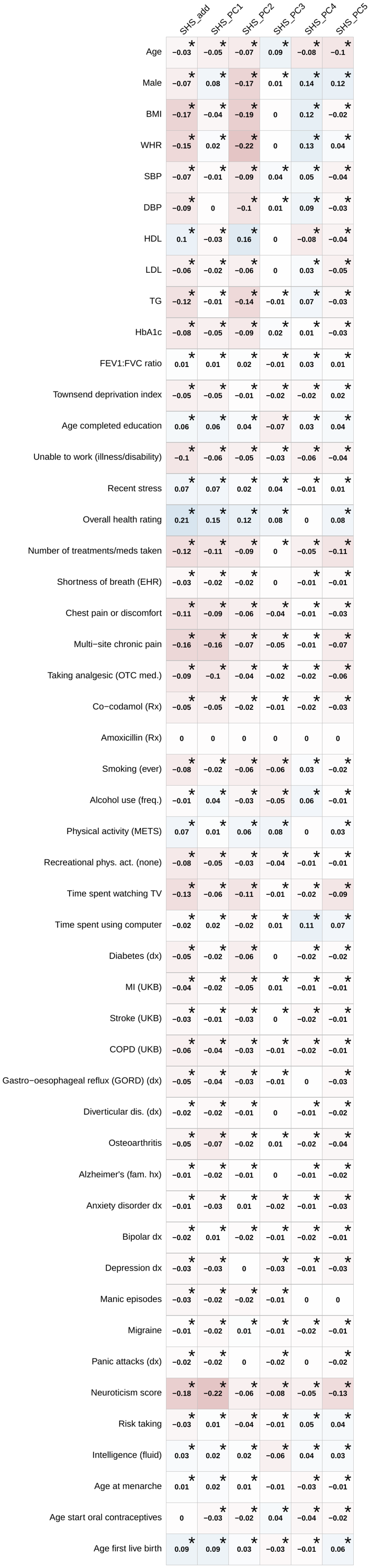

Health factors

General health & SES

Rx

Behavioral

Comorbid conditions

Psych. & cognitive

Reproductive

b. Genetic correlations with phenotypic traits of interest from LD score regression (*: p<2.2e-05).

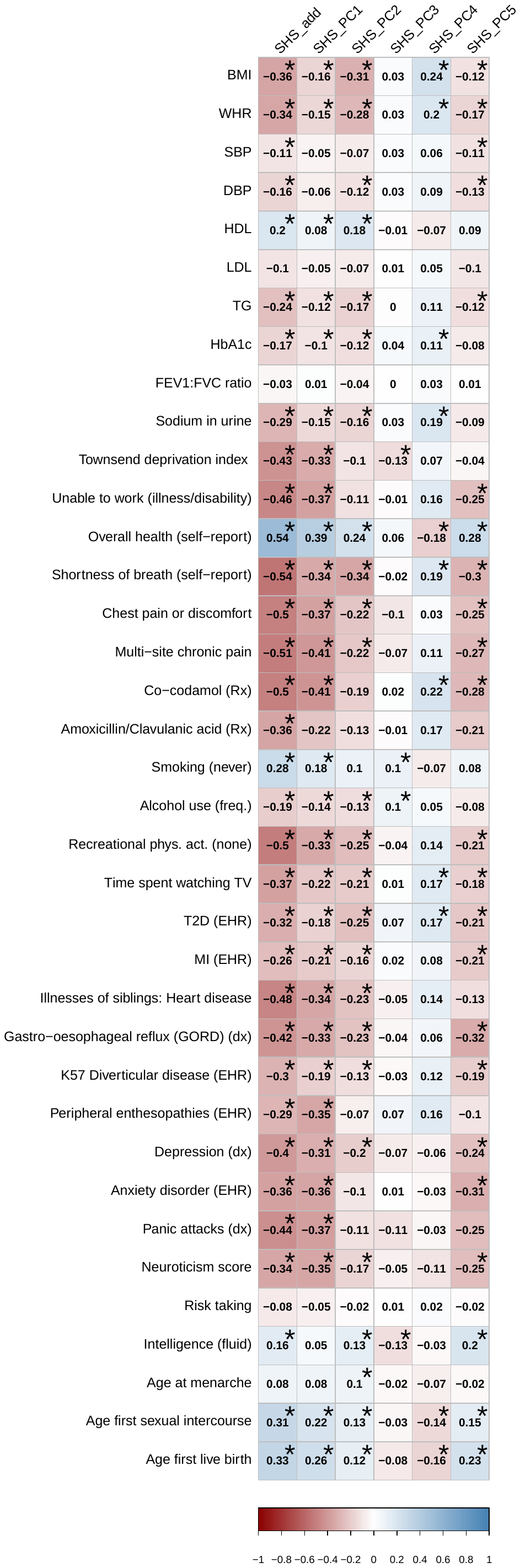

Health factors

General health & SES

Rx

Behavioral

Comorbid conditions

Psych. & cognitive

Reproductive

Supplementary Fig. 4. Circos Manhattan plot for SHS traits. The outer layer shows a Manhattan plot containing the negative log10-transformed P value of each SNP in the GWAS. Genome-wide significant SNPs (p<5e-8) are highlighted in blue. Genes were mapped in FUMA by position, chromatin interaction (black color in the inner circle) or eQTL (red color in inner circle).

a. SHS-ADD

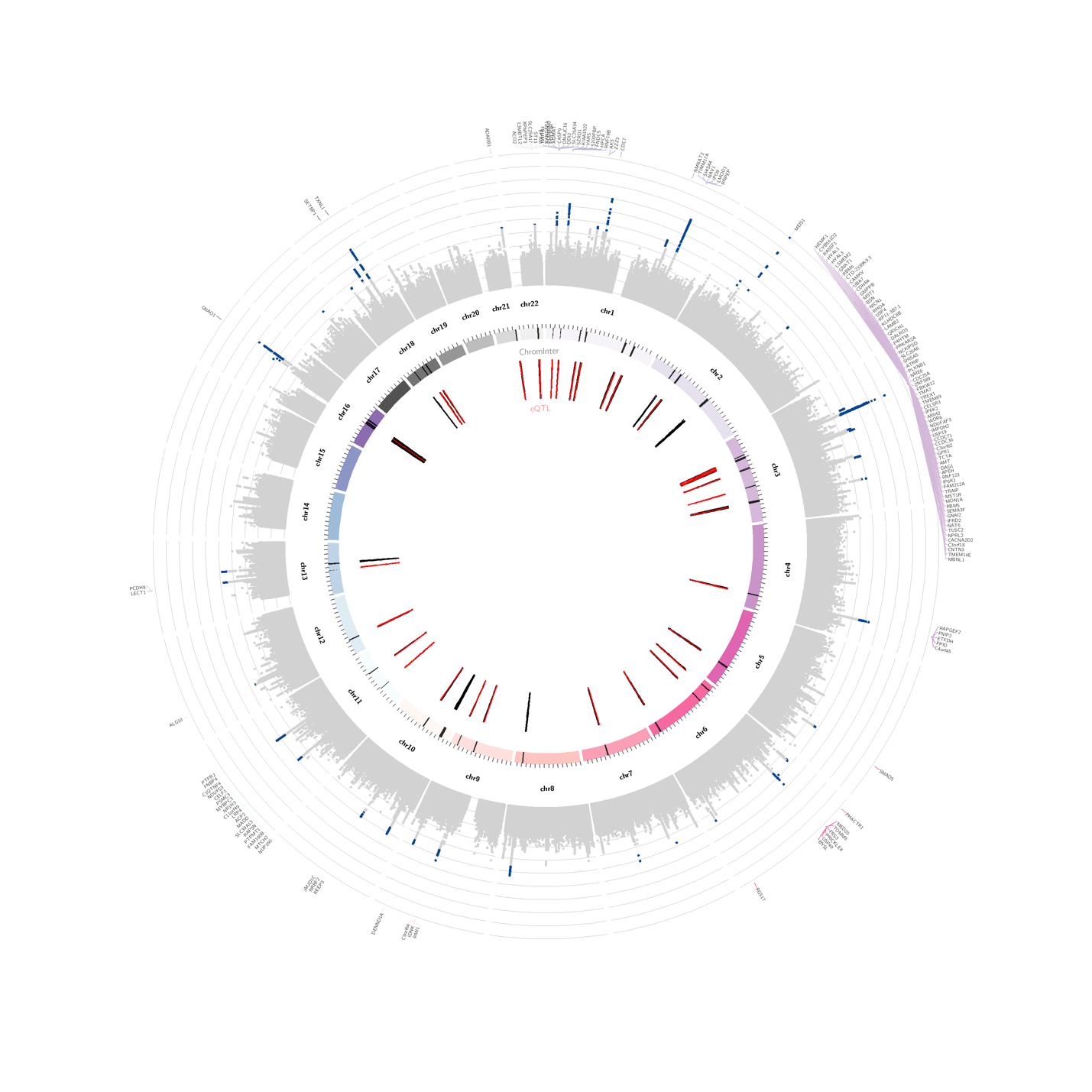

b. SHS-PC1

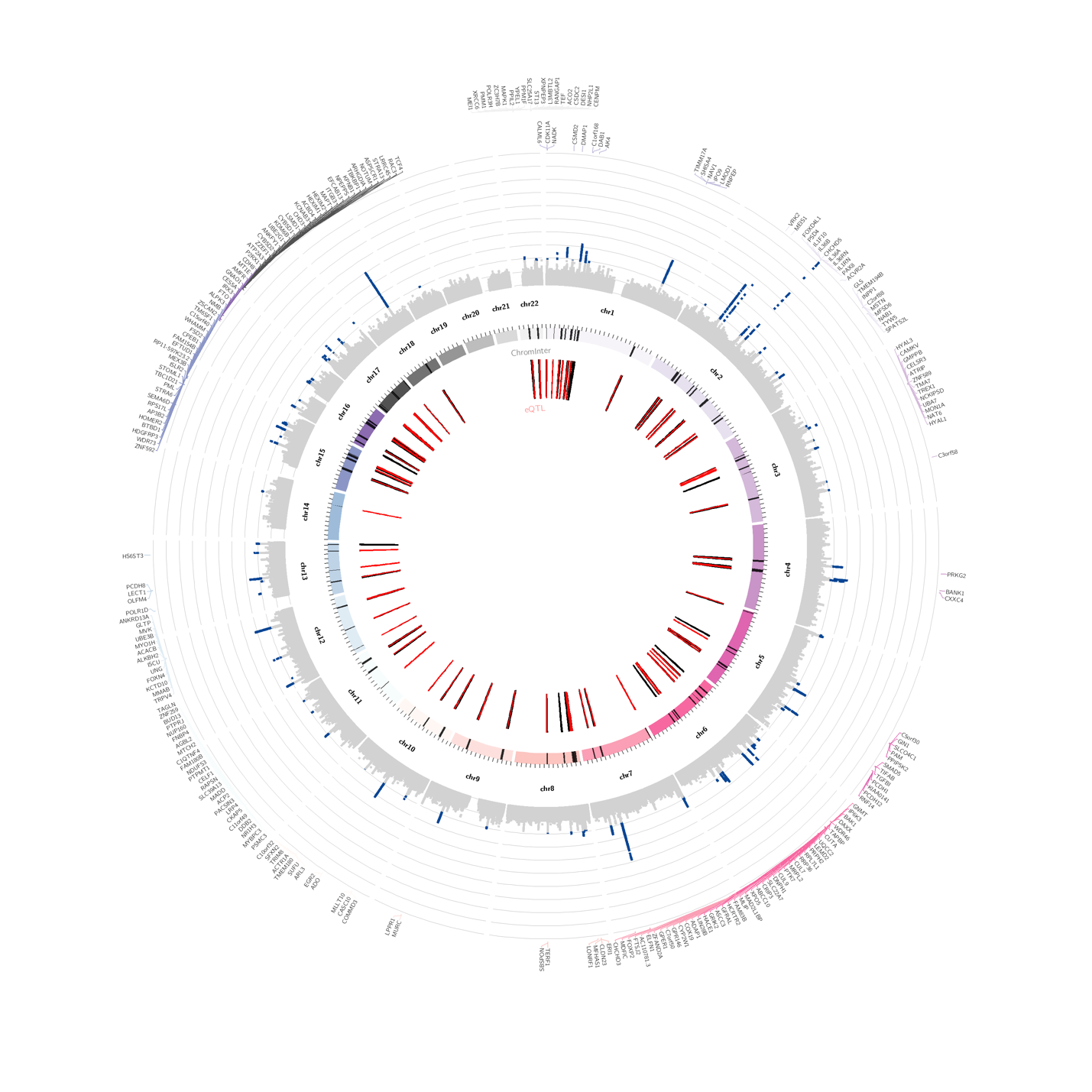

c. SHS-PC2

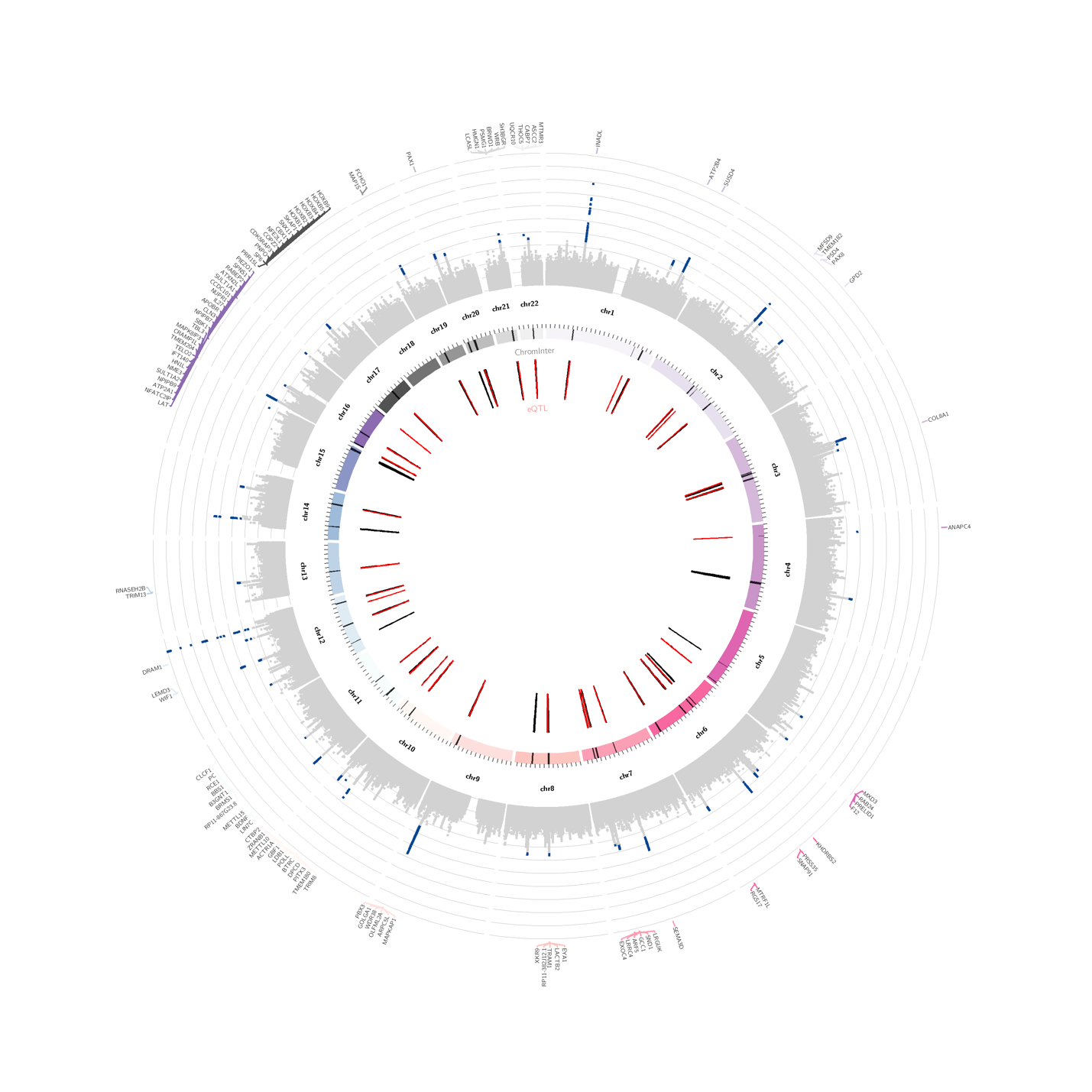

d. SHS-PC3

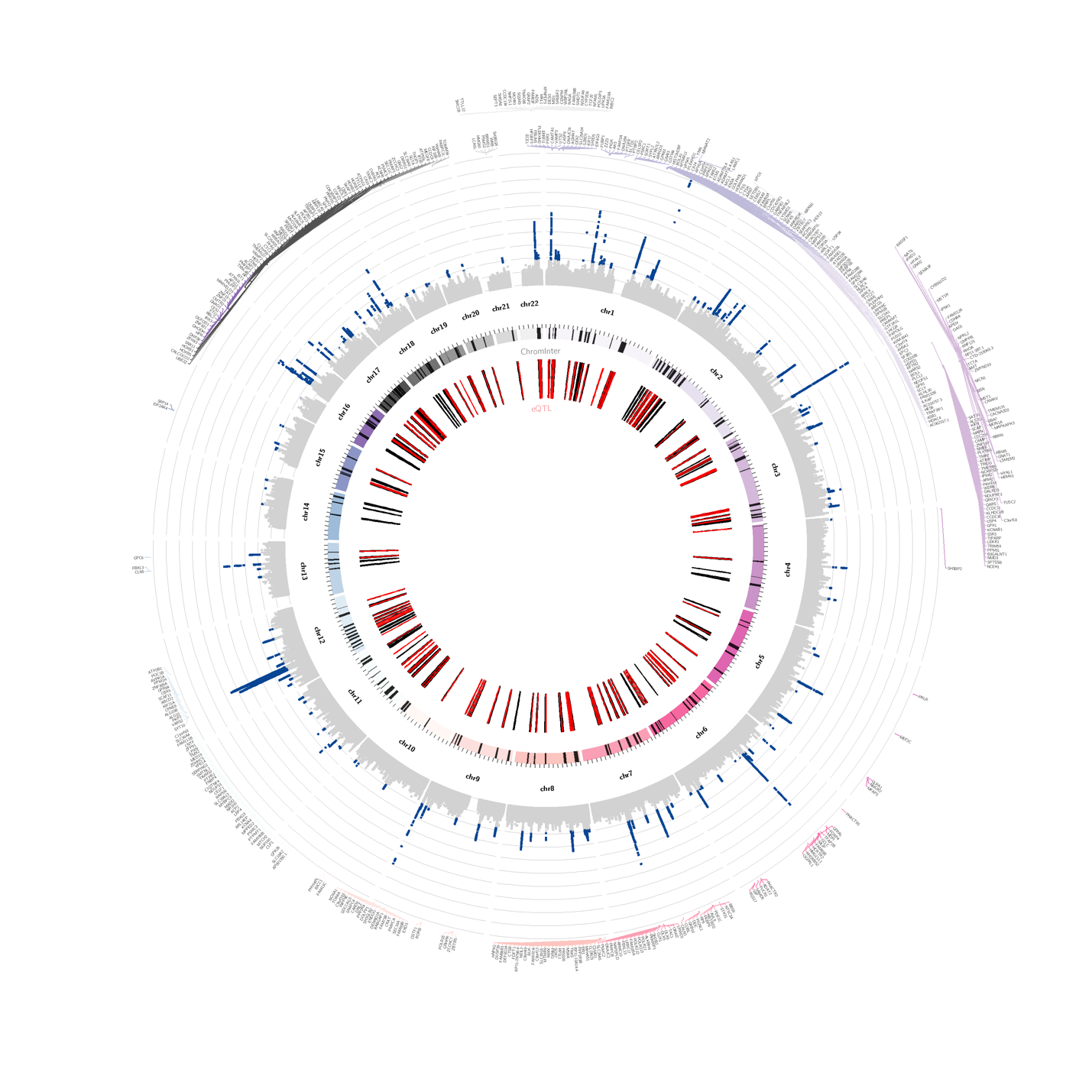

e. SHS-PC4

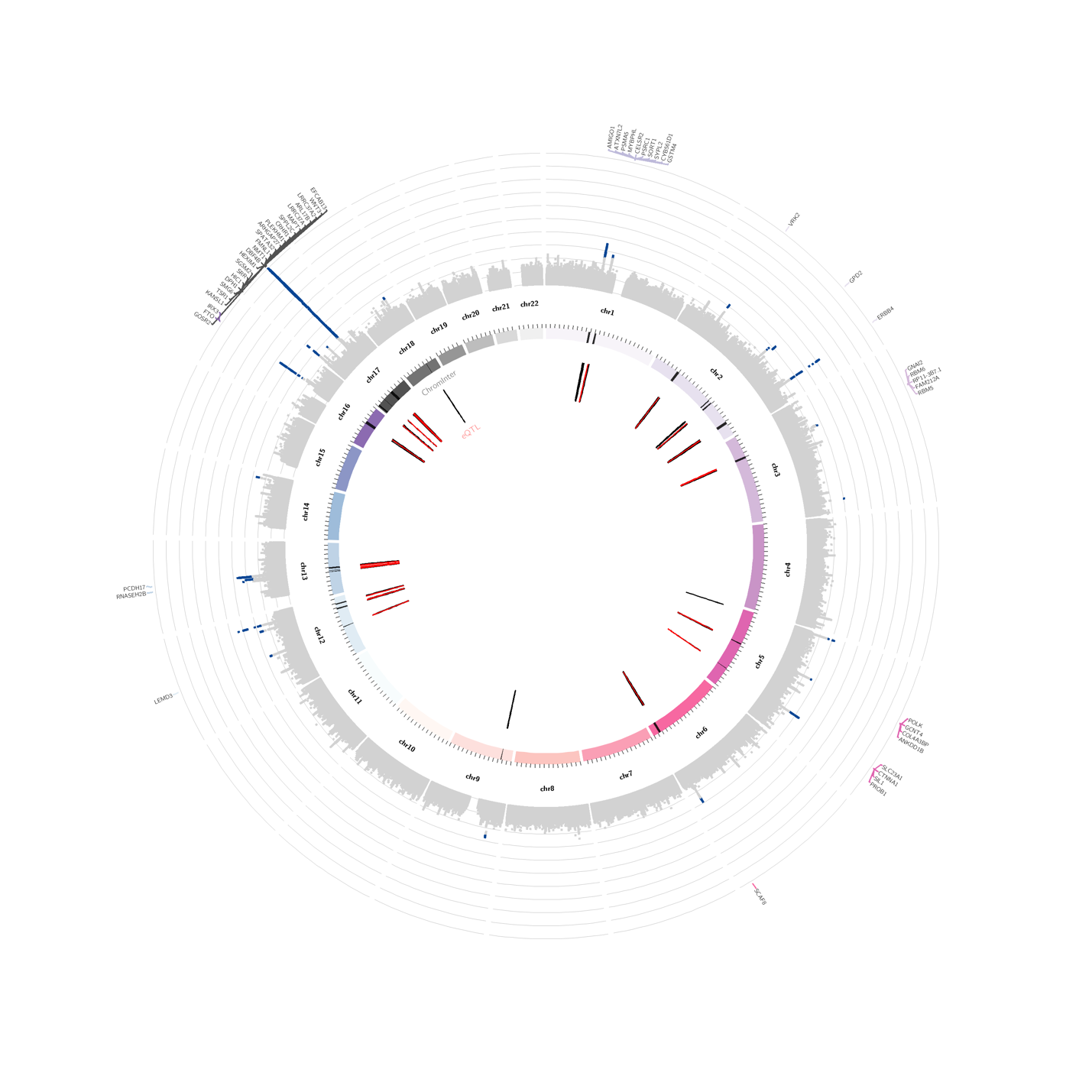

f. SHS-PC5

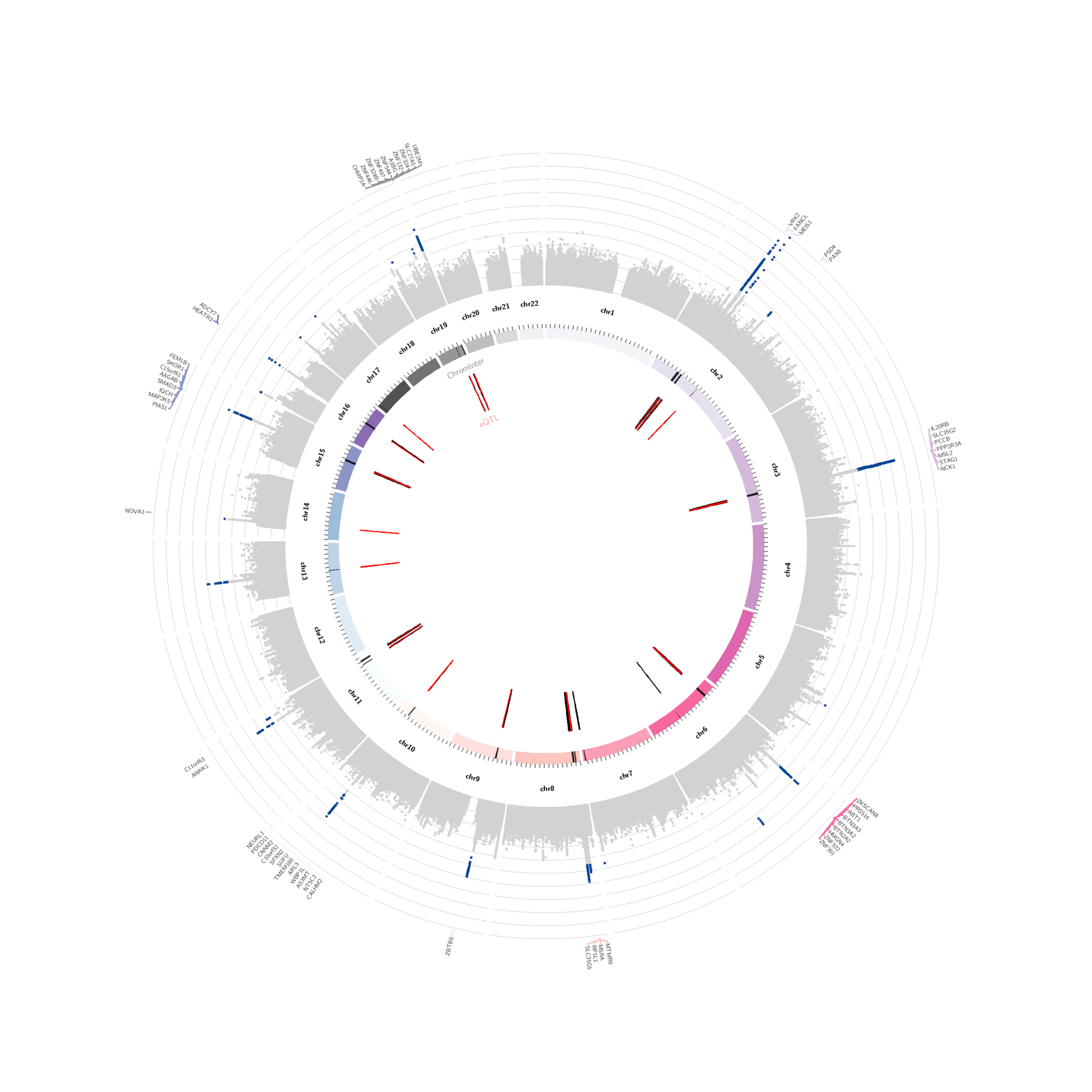

### Supplementary Fig. 5. QQ plots for SHS GWASs.

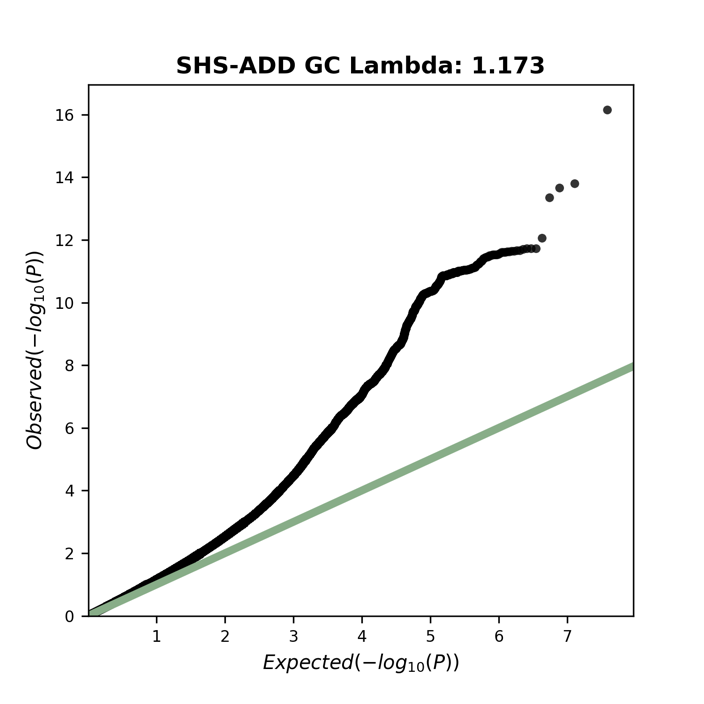

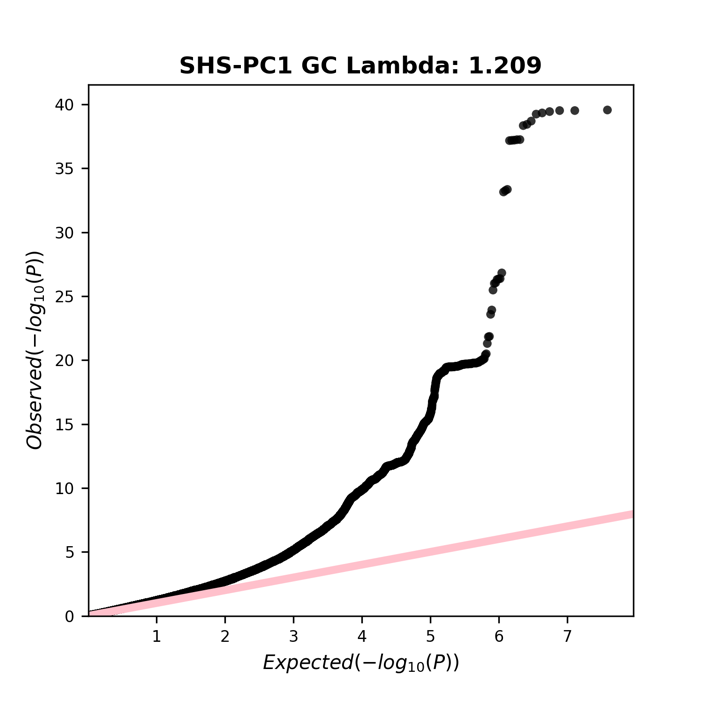

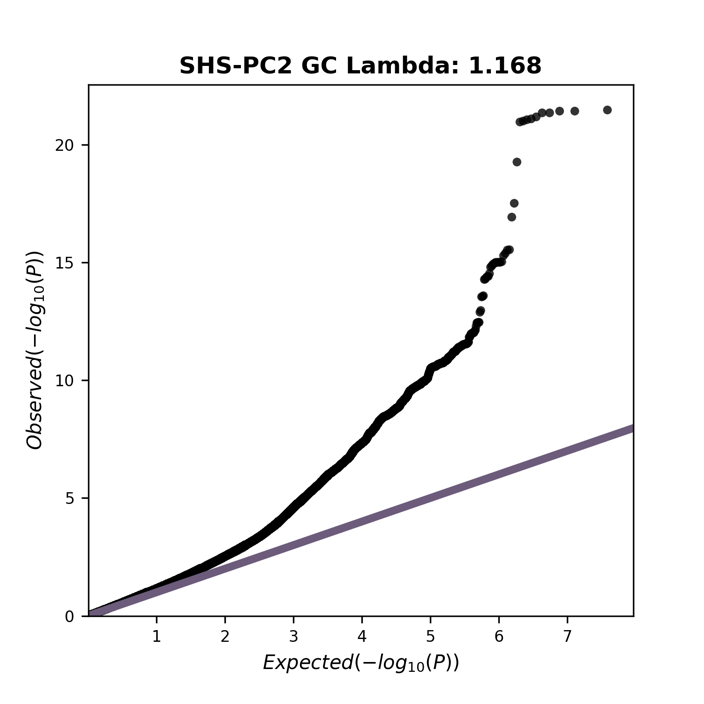

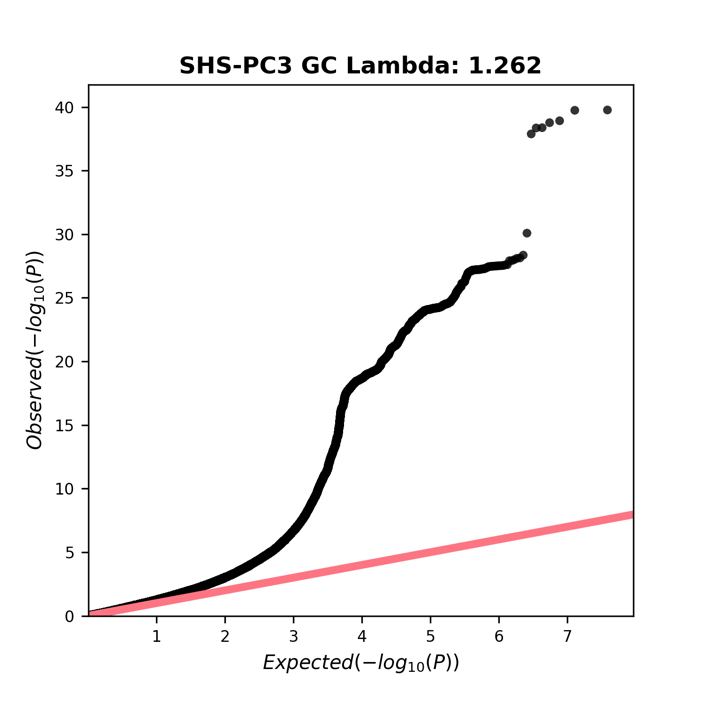

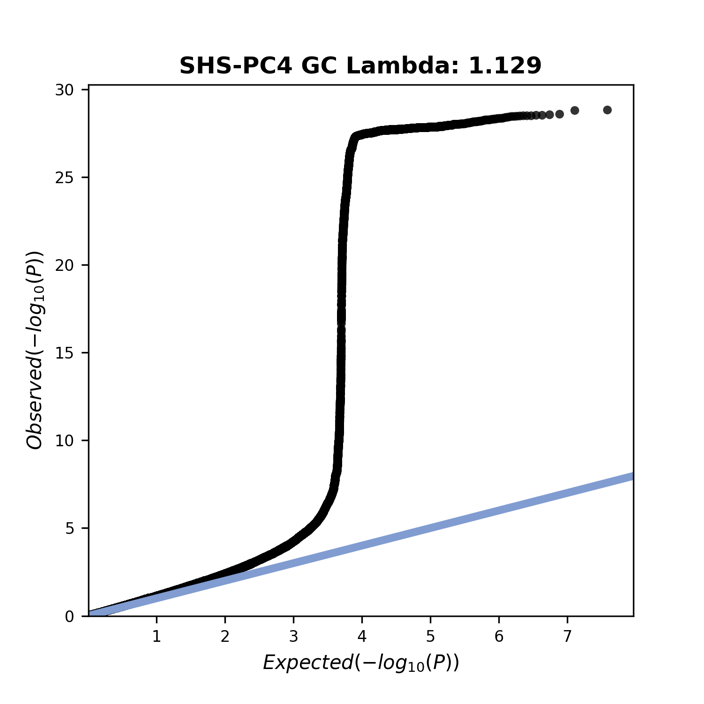

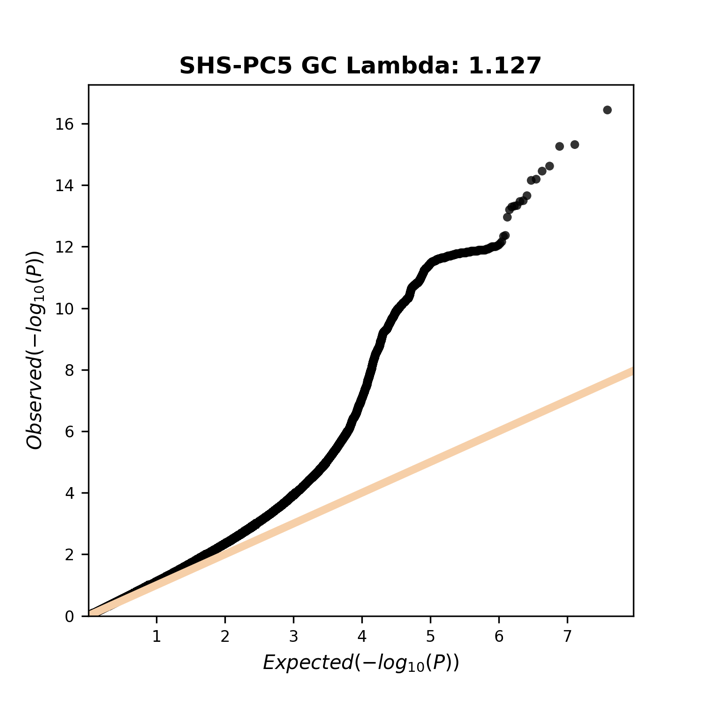

Supplementary Fig. 6. Manhattan plots of genome-wide gene-based analysis using FUMA MAGMA. Top 20 genes are labeled.

a. SHS-ADD

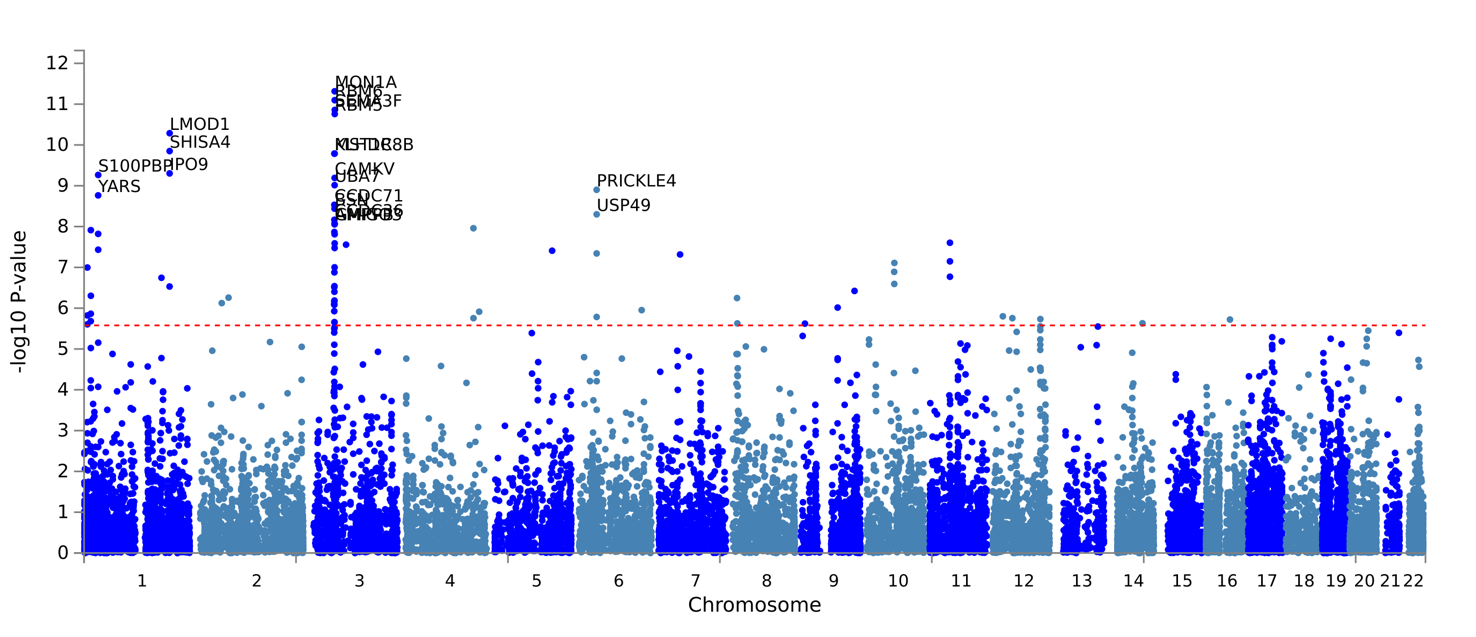

b. SHS-PC1

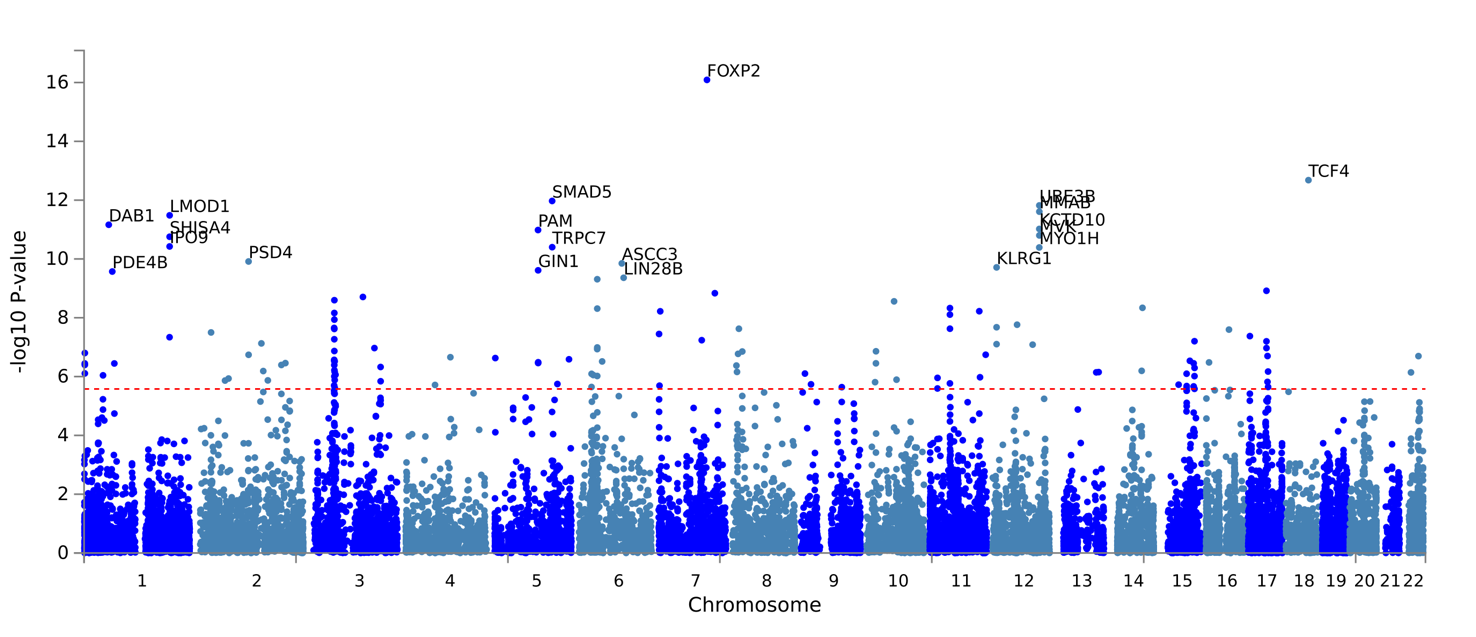

c. SHS-PC2

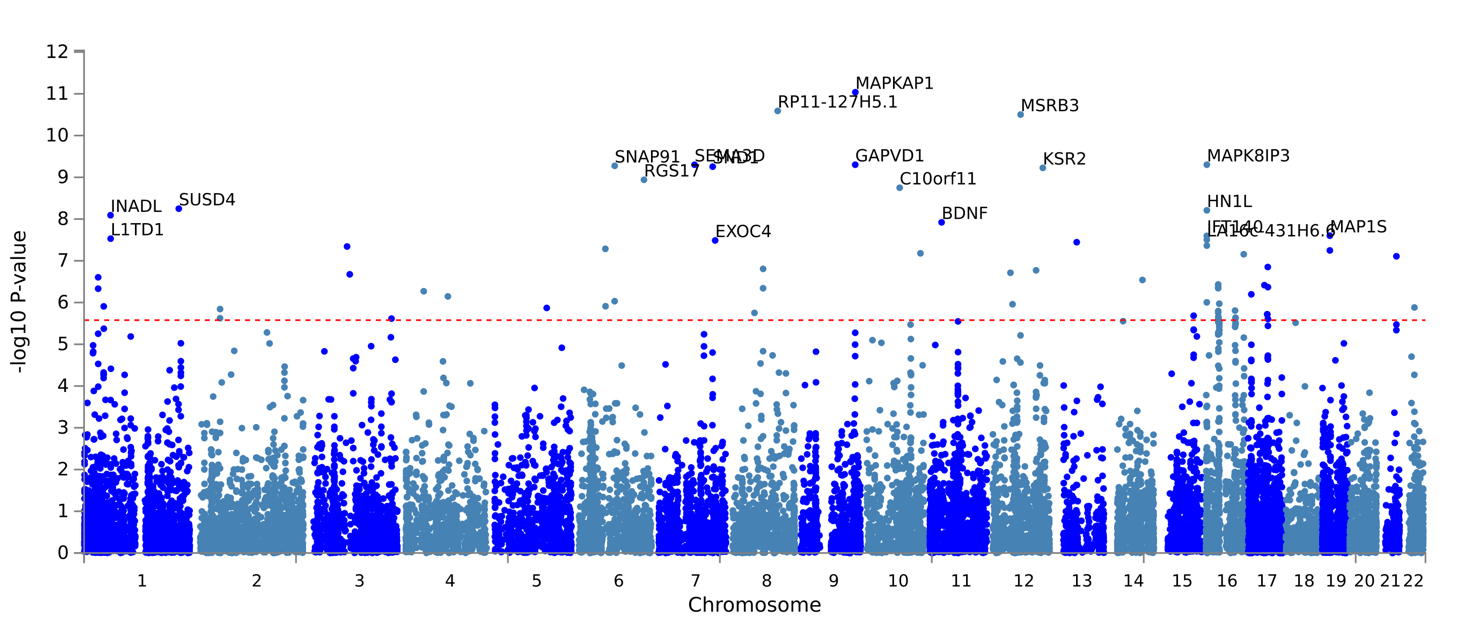

d. SHS-PC3

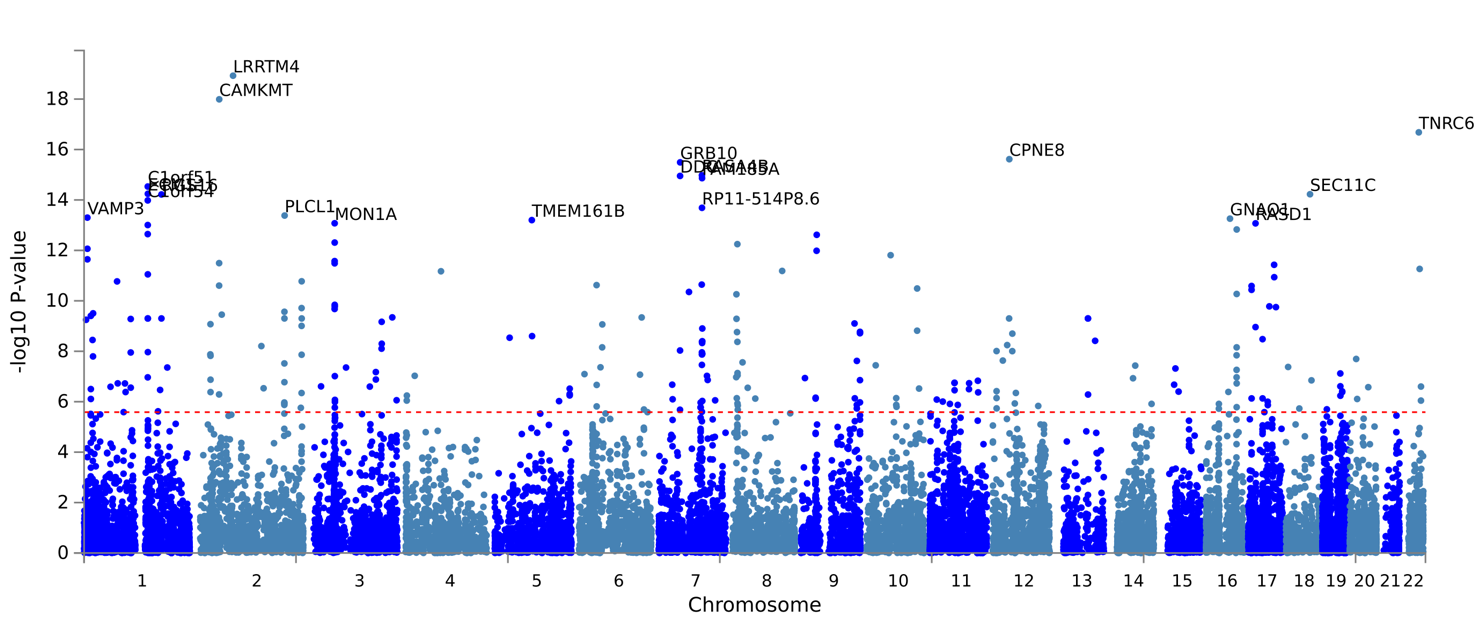

e. SHS-PC4
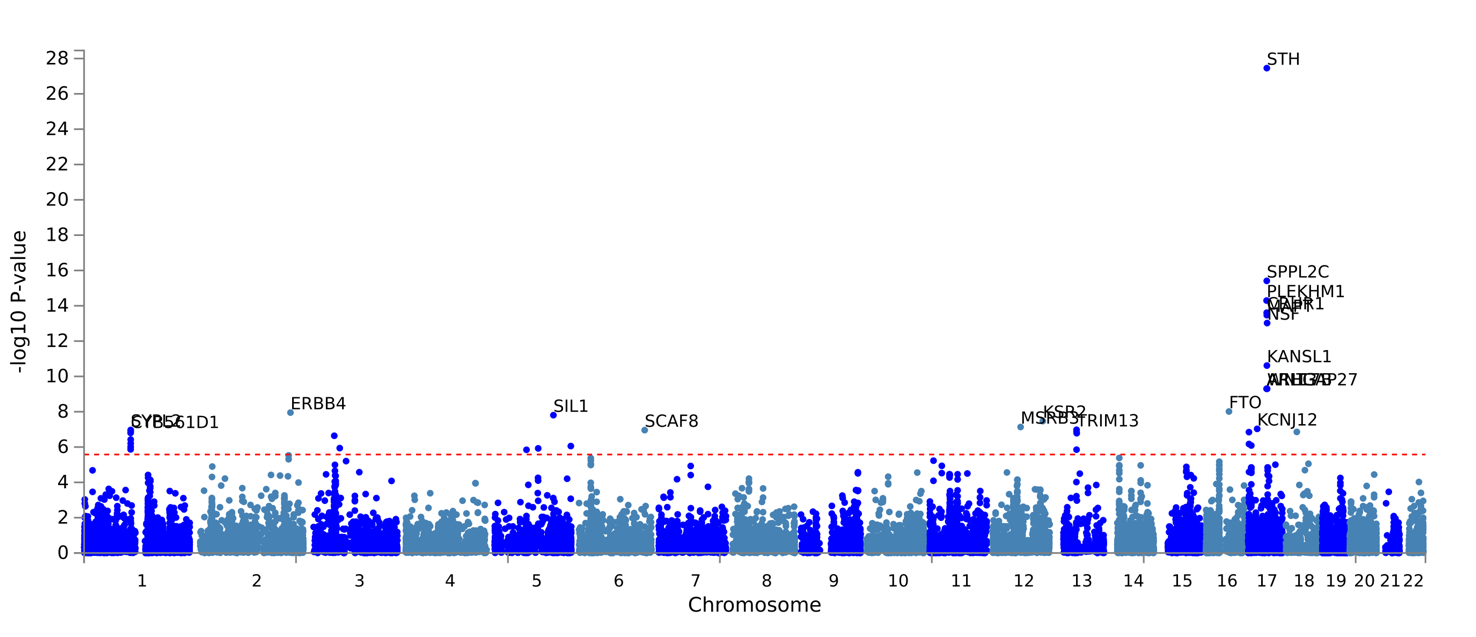

f. SHS-PC5

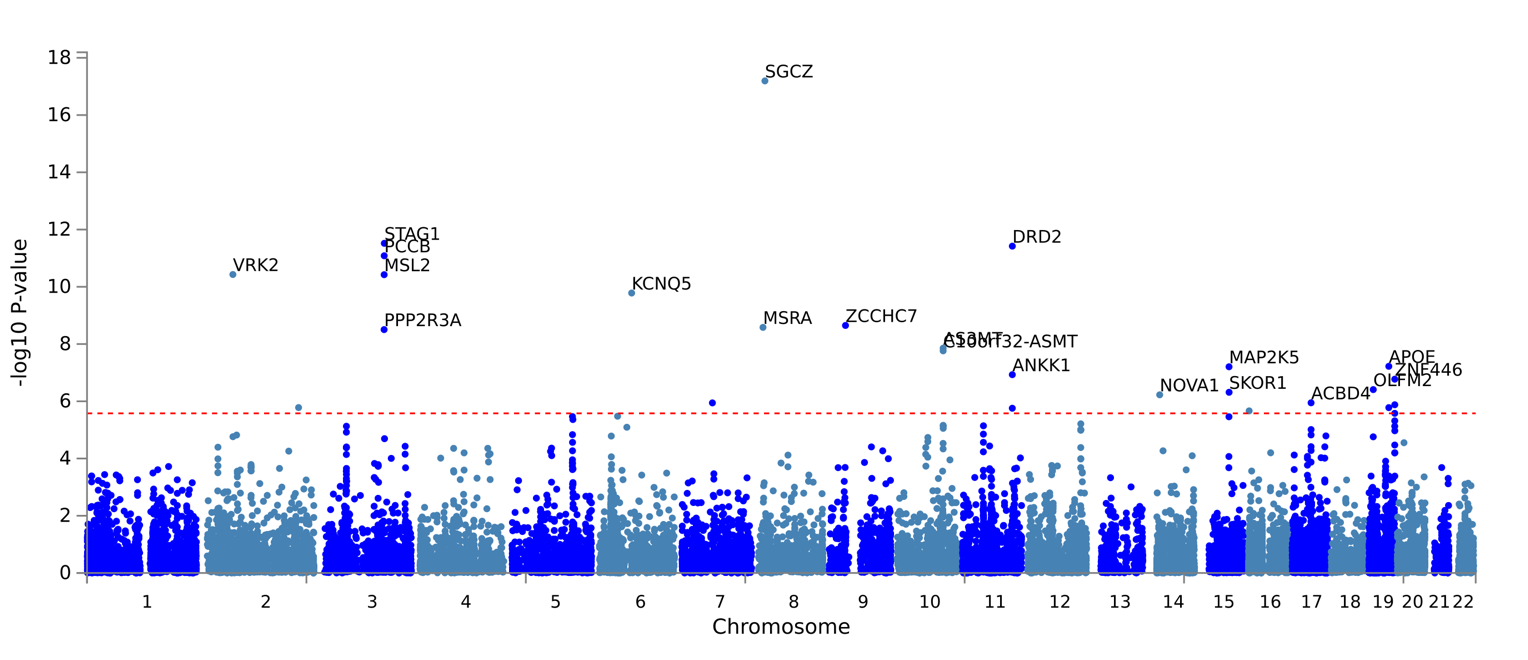

### Supplementary Fig. 7. QQ plots of genome-wide gene-based analysis using FUMA MAGMA.

| SHS-ADD  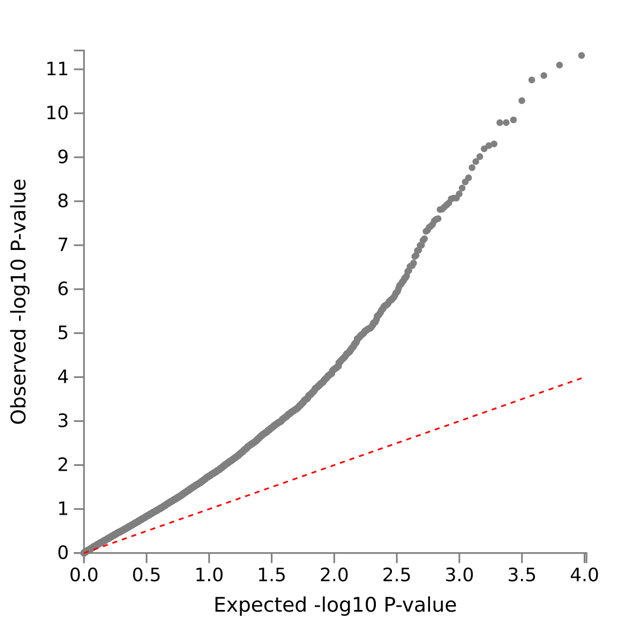 | SHS-PC1  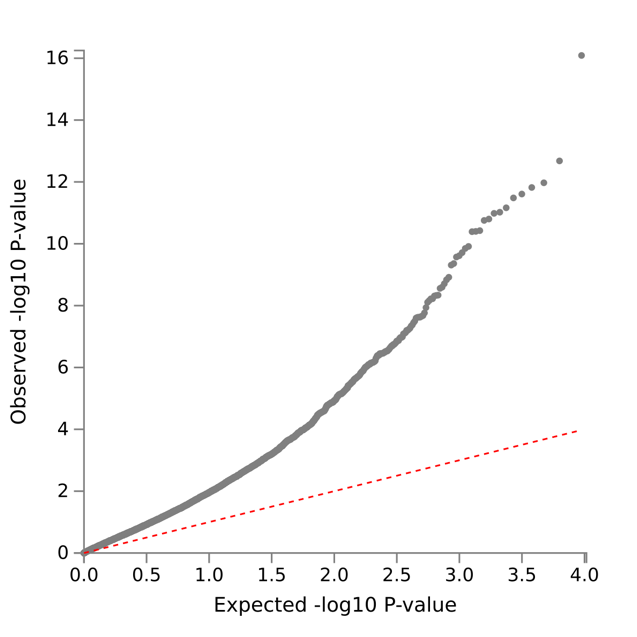 |
| --- | --- |
| SHS-PC2  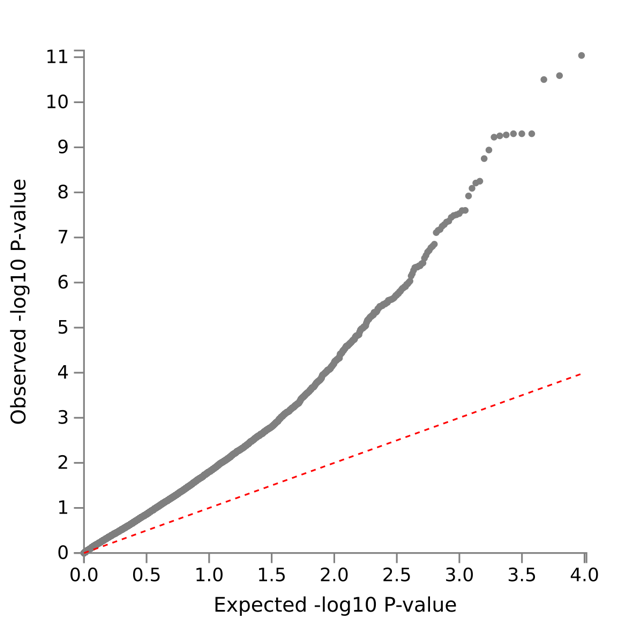 | SHS-PC3  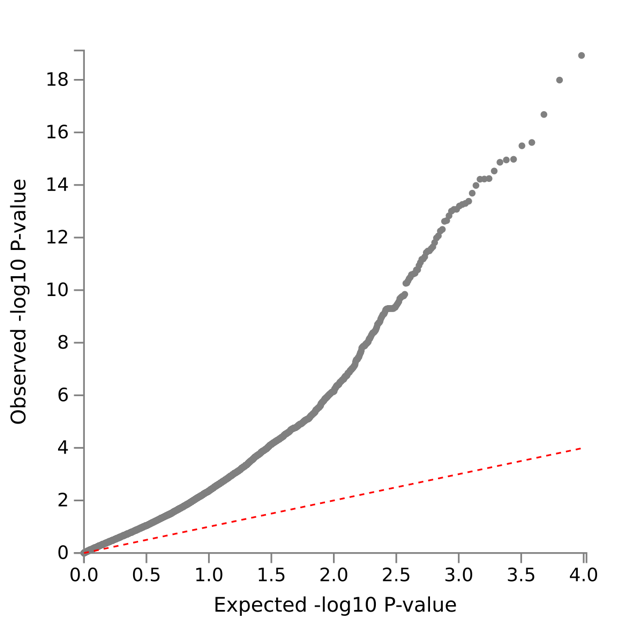 |
| SHS-PC4  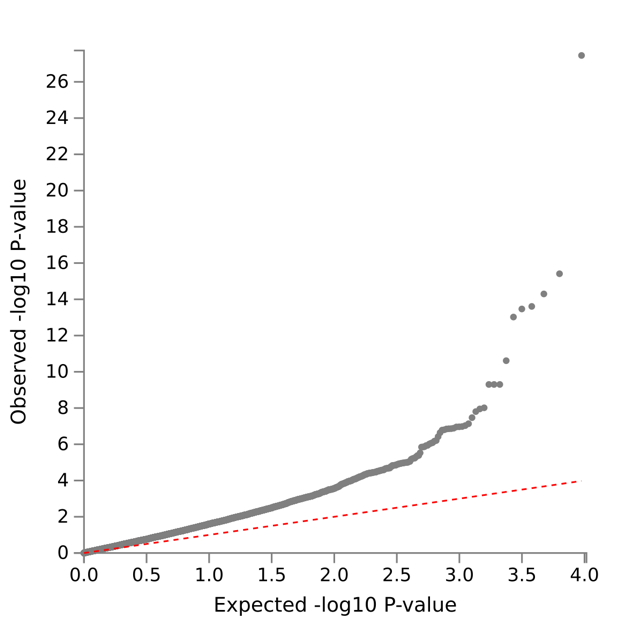 | SHS-PC5  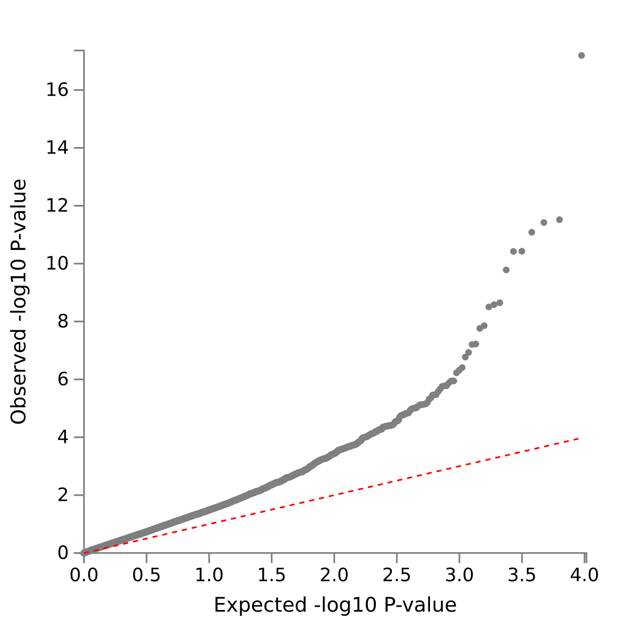 |

### Supplementary Fig. 8. Sensitivity results.

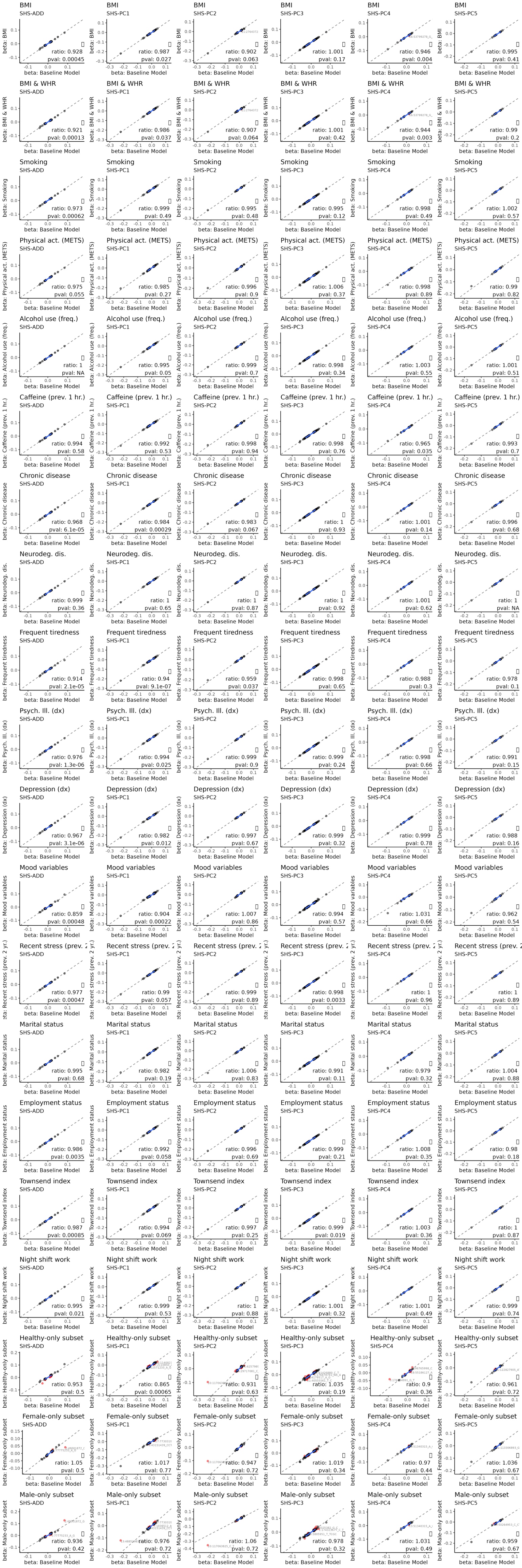

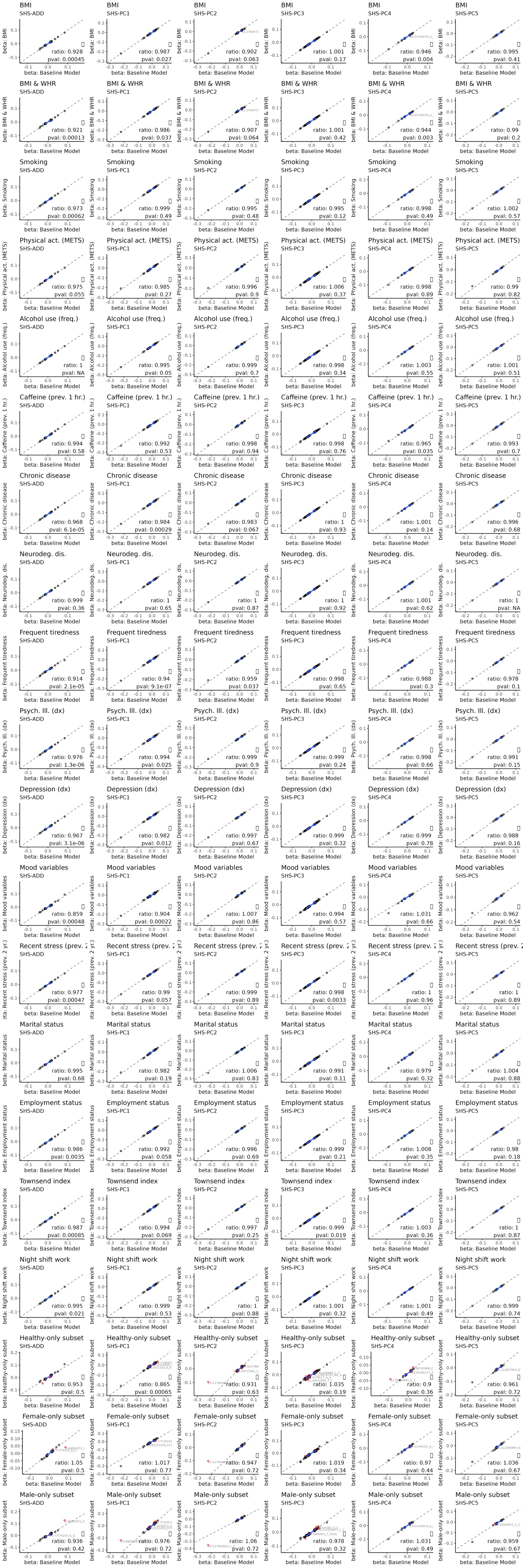

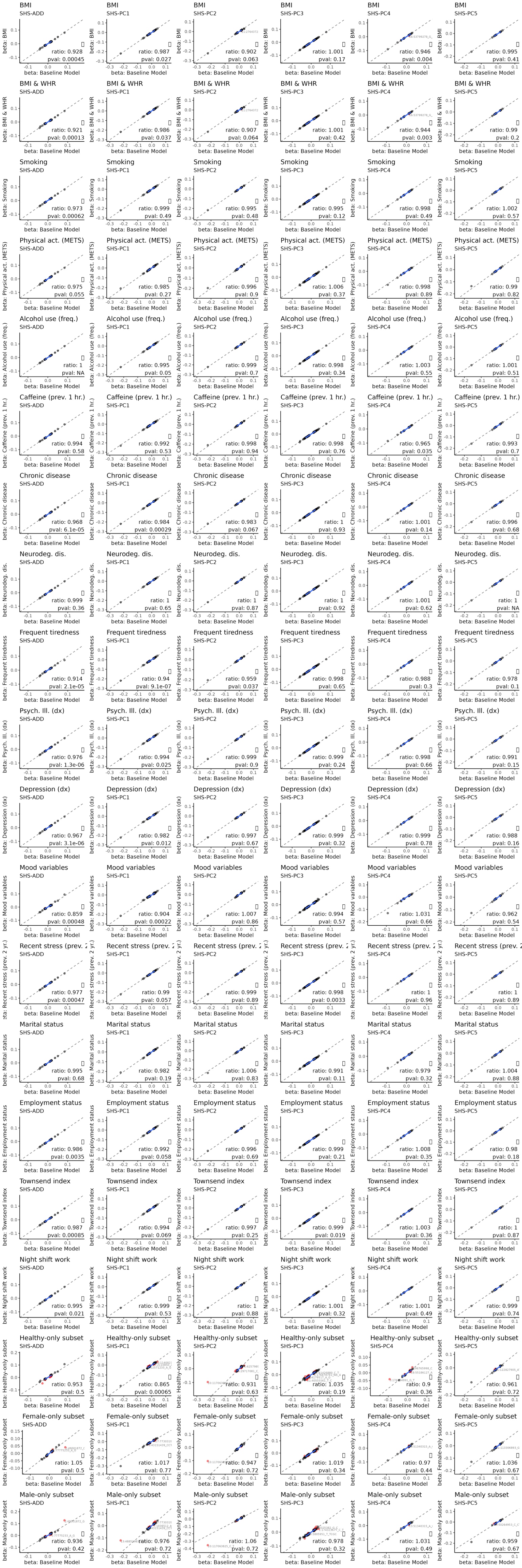

Plots of genetic effect size beta (log odds ratio) comparing baseline model with sensitivity models controlling for additional covariates. Corresponding variant-level results shown in Supplementary Table 12. Ratio: 90% truncated mean ratio of betas from sensitivity model compared with betas from baseline model. P-value: p-value for null hypothesis H_0_: Ratio = 1. Variants shown in red have adjusted effect size in the sensitivity model that differs from the baseline model by at least two standard errors.

### Supplementary Fig. 9. Significant cell types enriched for SHS genes in expression.

a. SHS-ADD

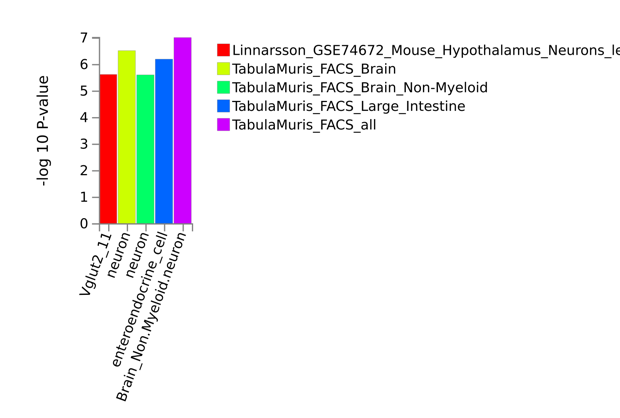

b. SHS-PC1

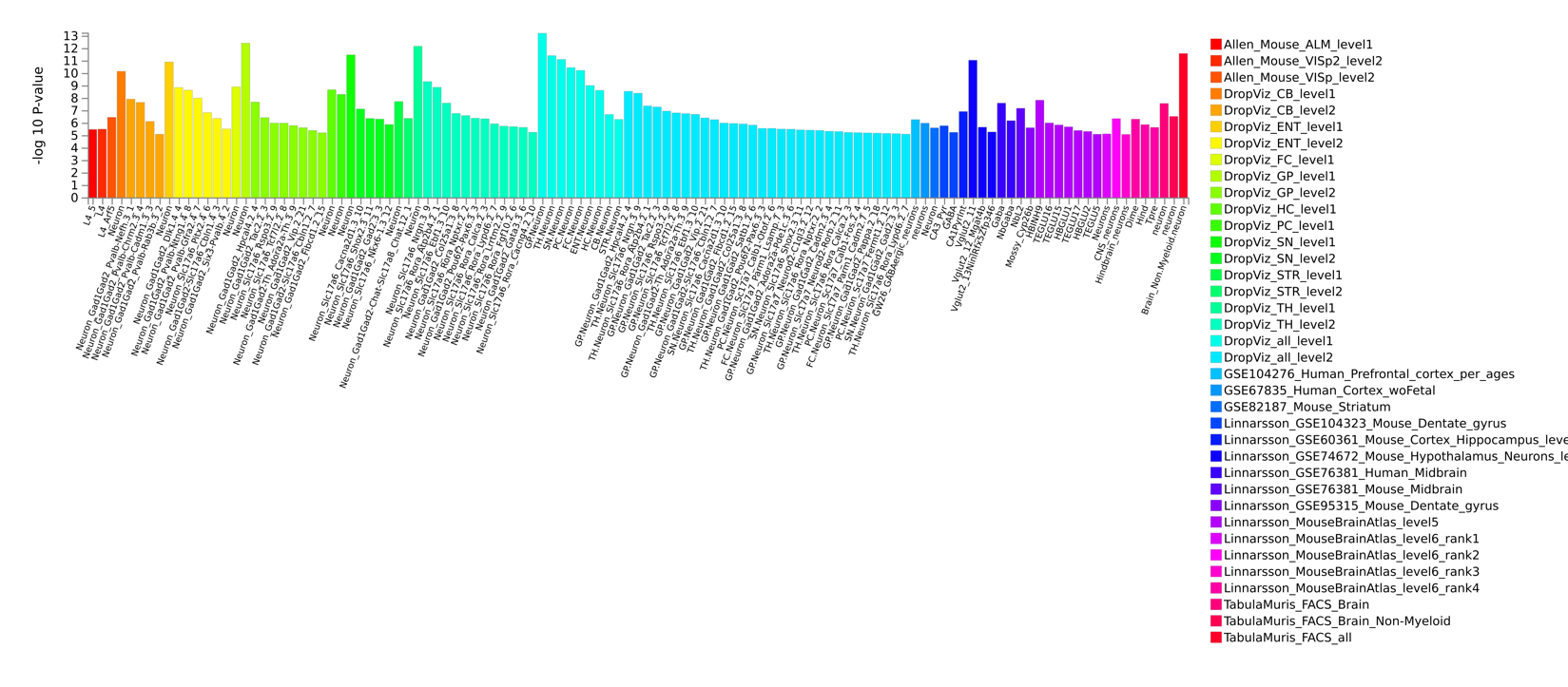

c. SHS-PC2

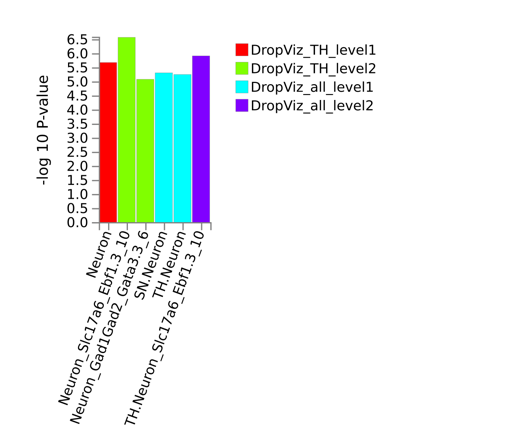

d. SHS-PC3

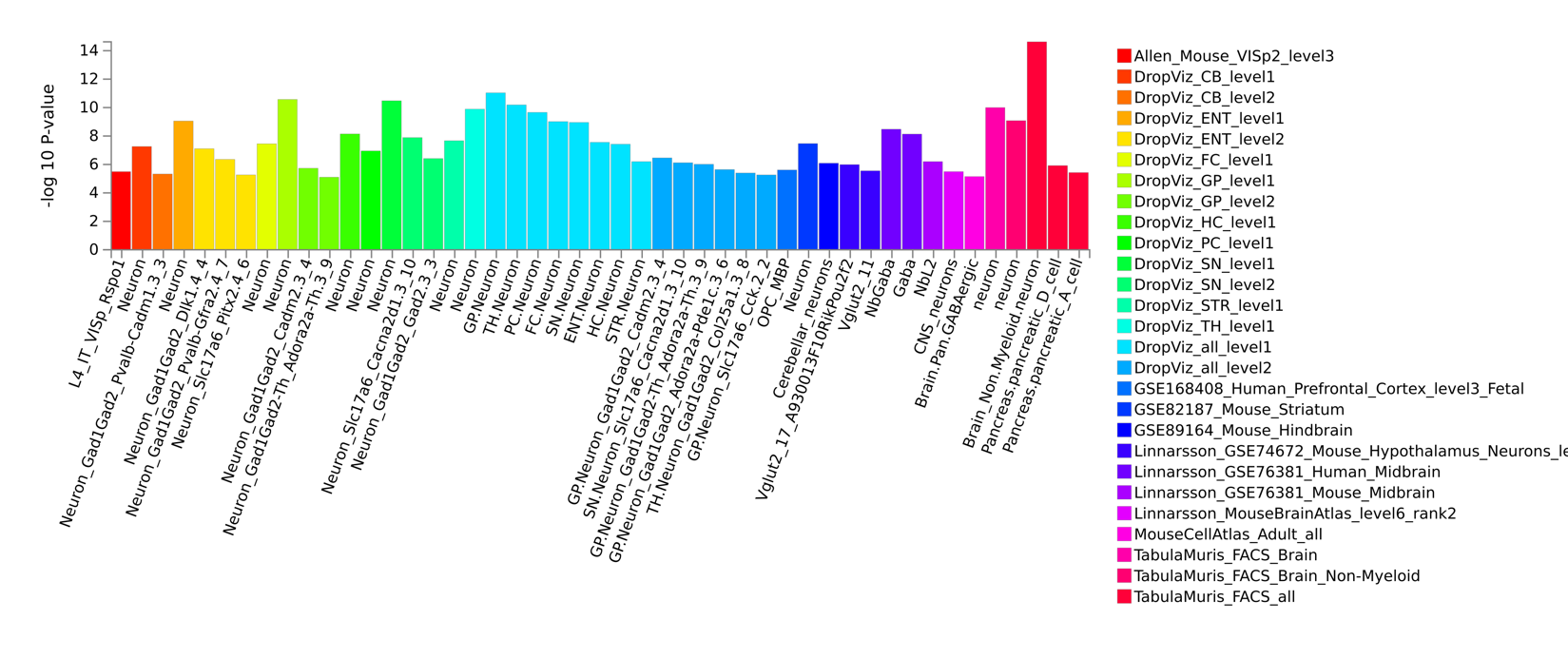

e. SHS-PC4

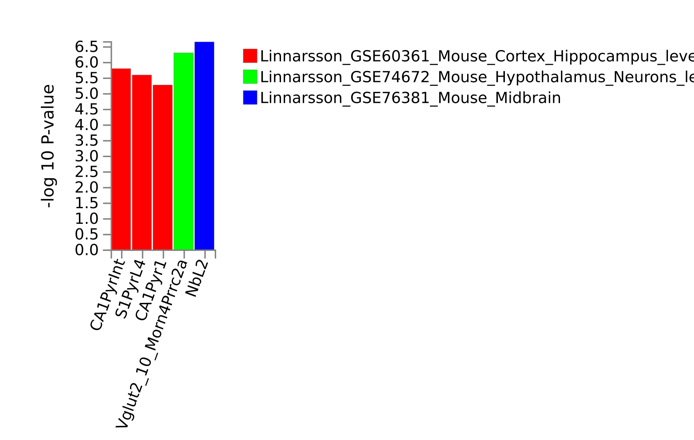

f. SHS-PC5

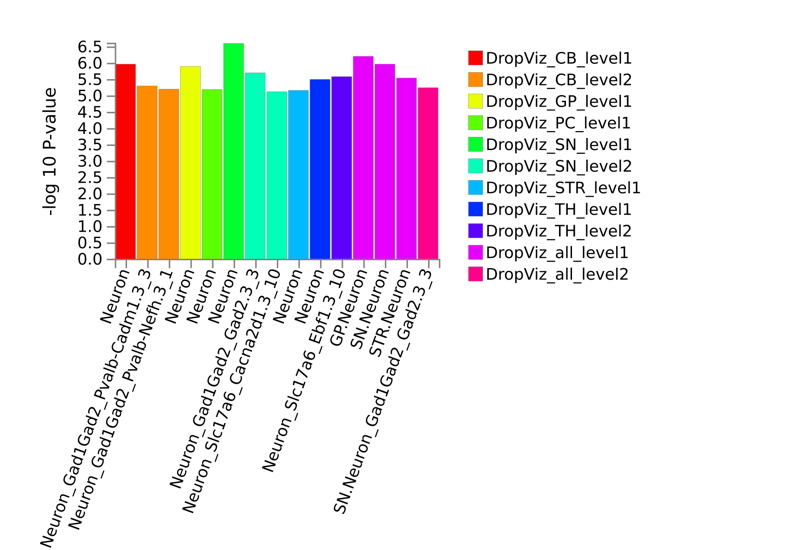
